# Health and Economic Effects of COVID-19 control in Australia: Modelling and quantifying the payoffs of ‘hard’ versus ‘soft’ lockdown

**DOI:** 10.1101/2020.08.31.20185587

**Authors:** R. Quentin Grafton, John Parslow, Tom Kompas, Kathryn Glass, Emily Banks, Kamalini Lokuge

## Abstract

**Objective(s):** Australia requires high quality evidence to optimise likely health and economy outcomes to effectively manage the current resurgence of COVID-19. We hypothesise that the most stringent social distancing (SD) measures (100% of level in Australia in April 2020) deliver better public health and economy outcomes.

**Design:** ‘Fit-for-purpose’ (individual-based and compartment) models were used to simulate the effects of different SD and detection strategies on Australian COVID-19 infections and the economy from March to July 2020. Public reported COVID-19 data were used to estimate model parameters.

**Main outcome measures:** Public health and economy outcomes for multiple social distancing levels were evaluated, assessing “hard” versus “soft” lockdowns, and for early versus later relaxation of social distancing. Outcomes included costs and the timing and magnitude of observed COVID-19 cases and cumulative deaths in Australia from March to June 2020.

**Results:** Higher levels of social distancing achieve zero community transmission with 100% probability and lower economy cost while low levels of social distancing result in uncontrolled outbreaks and higher economy costs. High social distancing total economy costs were $17.4B versus $41.2B for 0.7 social distancing. Early relaxation of suppression results in worse public health outcomes and higher economy costs.

**Conclusion(s):** Better public health outcomes (reduced COVID-19 fatalities) are *positively* associated with lower economy costs and higher levels of social distancing; achieving zero community transmission *lowers* both public health and economy costs compared to allowing community transmission to continue; and early relaxation of social distancing *increases* both public health and economy costs.

**Significance:** *The known* is that COVID-19 infections can be suppressed with social distancing (SD) measures of sufficient stringency and duration.

*The new* is we find highest levels of SD (100% SD that prevailed in April 2020) generate much lower COVID-9 deaths; reduced SD days; increased economic activity; and much higher probability of elimination over a subsequent 12-month period than lower levels of SD.

*The implications* are that greater levels of SD are preferred to lower SD because they deliver both better public health and lower economy costs.

## Manuscript

### 1. Introduction

Australia recorded its first case of COVID-19 on 25 January 2020 from a person who had flown from China on 19 January^1^. On 6 August 2020, there were: approximately 20,000 reported cumulative cases, of which some 11,000 had recovered; in total, about 250 COVID-19 fatalities (162 in Victoria); approximately 7,500 active cases; and 739 additional recorded cases nationally over the previous 24 hours; almost all in Victoria (725 cases).^2^

Fig. 1 shows the cumulative number of reported Australian cases and current cases (infected less recovered less fatalities) from 26 March to 6 August 2020. Initial daily infections peaked at 458 recorded cases on 28 March and declined due to border measures for overseas arrivals, self-quarantine, wide-spread testing and contact tracing, and social distancing outside of households that changed the frequency, number and nature of physical contacts. Combined, these measures were highly successful such that by 9 June there were only 2 new recorded cases in Australia. From mid-June onwards, outbreaks of COVID-19 were recorded in Victoria or associated with persons who were infected in Victoria.

**Fig. 1:**
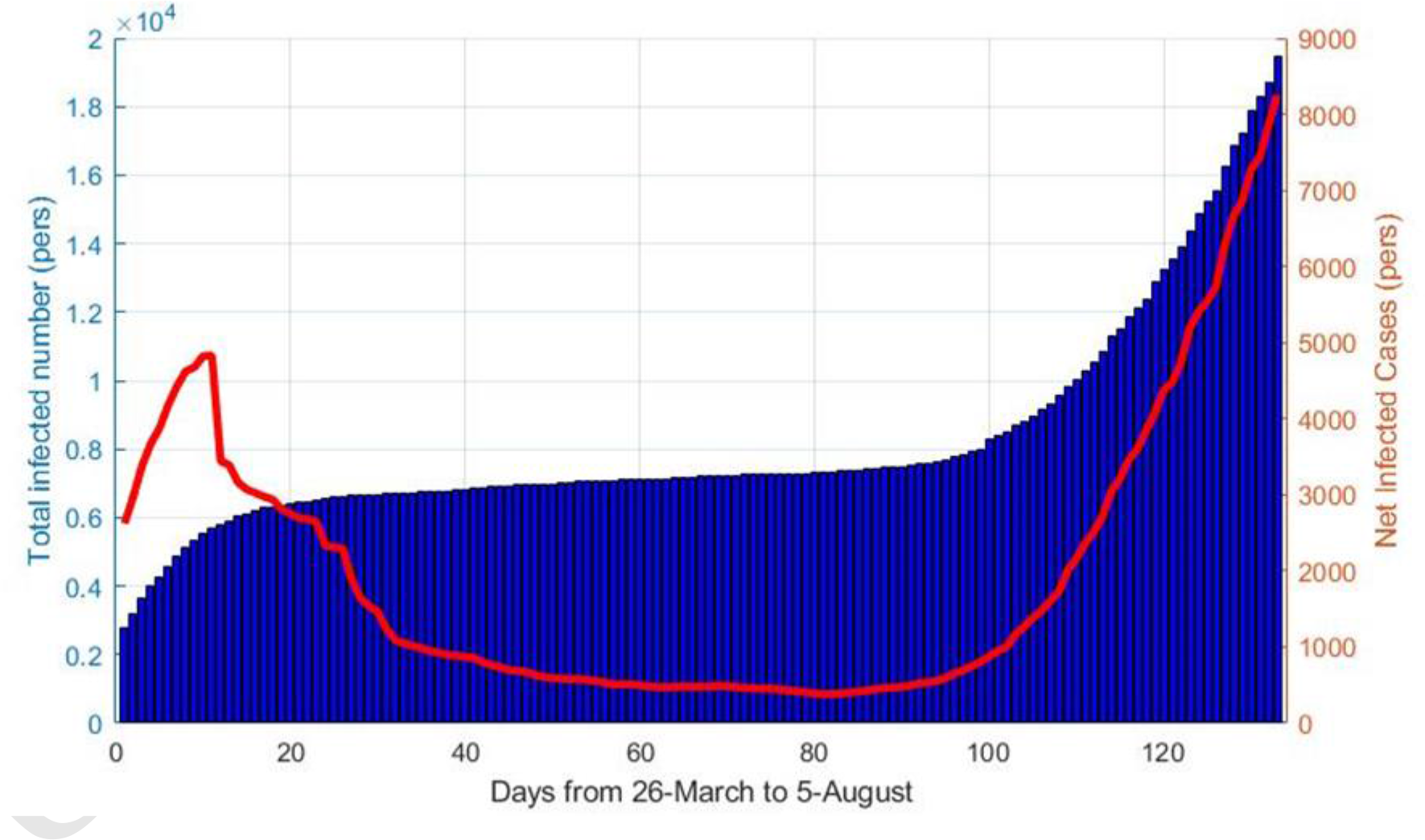
Cumulative (total infected) and current (net infected) reported COVID-19 cases in Australia from March 26 to 6 August 2020

Public health responses to COVID-19 have evolved over time. From February to March 2020, a series of control measures were implemented at a national level beginning with travel restrictions from China to Australia on 1 February, followed by a requirement that all international arrivals self-quarantine for 14 days from 15 March. On 18 March, non-essential gatherings were banned and individual physical distancing encouraged (1.5 m distance between people and 4 m^2^ per person), on 21 March non-essential businesses were closed, and on 30 March public gatherings were restricted to a maximum of two people^3^. These measures successfully reduced infections^4^ such that on the 10 May, social distancing restrictions began to be relaxed and, by the end of June, the level of social distancing in most Australian jurisdictions^5^, including Victoria, was at near to pre-COVID-19 levels^6^.

Additional public health measures were reinstituted in the second half of June in Victoria following leakage from hotel quarantine in Melbourne in late May. Multiple outbreaks in Melbourne resulted in the introduction of a ‘stage 3’ lockdown in Melbourne and Mitchell Shire on 9 July for a 6-week period. Despite a massive campaign of testing and contact tracing, the reintroduction of mandated social distancing, and the mandated use of masks outside on 22 July, these measures failed to adequately reduce new COVID-19 infections. Nevertheless, combined these measures may have averted at least 9,000 additional COVID-19 cases in July^7^.

As a result of the resurgence of COVID-19 infections, the Victorian state government implemented a 8:00 pm to 5:00 am curfew in Melbourne that began 2 August and a 6-weeks ‘stage 4’ lockdown for Melbourne similar to what New Zealand implemented^8^ that began 6 August and which includes school closures and a requirement for all but essential workers to stay at home. These social distancing measures in Melbourne are complemented by a 6-weeks ‘stage 3’ lockdown for the rest of Victoria that also began 6 August 2020.

The key public health and economy questions facing Australia, and to which this study responds, are: What levels of lockdown (as measured by a social distancing level) are required to adequately reduce infections associated with the COVID-19 Victorian resurgence? What is the probability of achieving elimination (defined as no community transmission^9^) with various levels of lockdown and duration? What are the public health and economy costs of alternative levels of social distancing (corresponding to different severity levels of lockdowns)?

### 2. Methods

A suite of epidemiological models was developed to simulate the response of COVID-19 outbreaks to a range of control measures in Australia and Victoria. These models are designed to offer a flexible and efficient representation of the progression of the disease in individual cases, transmission of the disease within the susceptible population, and the effects of community testing, downstream contact tracing, self-isolation and quarantine on detection and transmission. The model suite comprises an individual-based model (IBM), a stochastic compartment model (SCM), and a deterministic compartment model (DCM). All three are numerical models that operate in discrete time with daily time steps.

#### Individual-based Model (IBM)

The IBM follows infected individuals through time, starting on the day when they are infected, and ending when they are officially recovered. Individuals are characterised by a set of attributes whose values evolve over time according to a set of rules. All attributes, with one exception, take logical values of zero or 1, and determine the individual’s status with respect to source of infection (i.e. local or overseas), infectivity, display of symptoms, detection, traceability, self-isolation, self-quarantine or quarantine, recovery, hospitalisation, and/or death. The attributes are listed in Table 1 of the supplementary material. The default values of all attributes are set to zero, until they are modified by model processes.

**Table 1.**
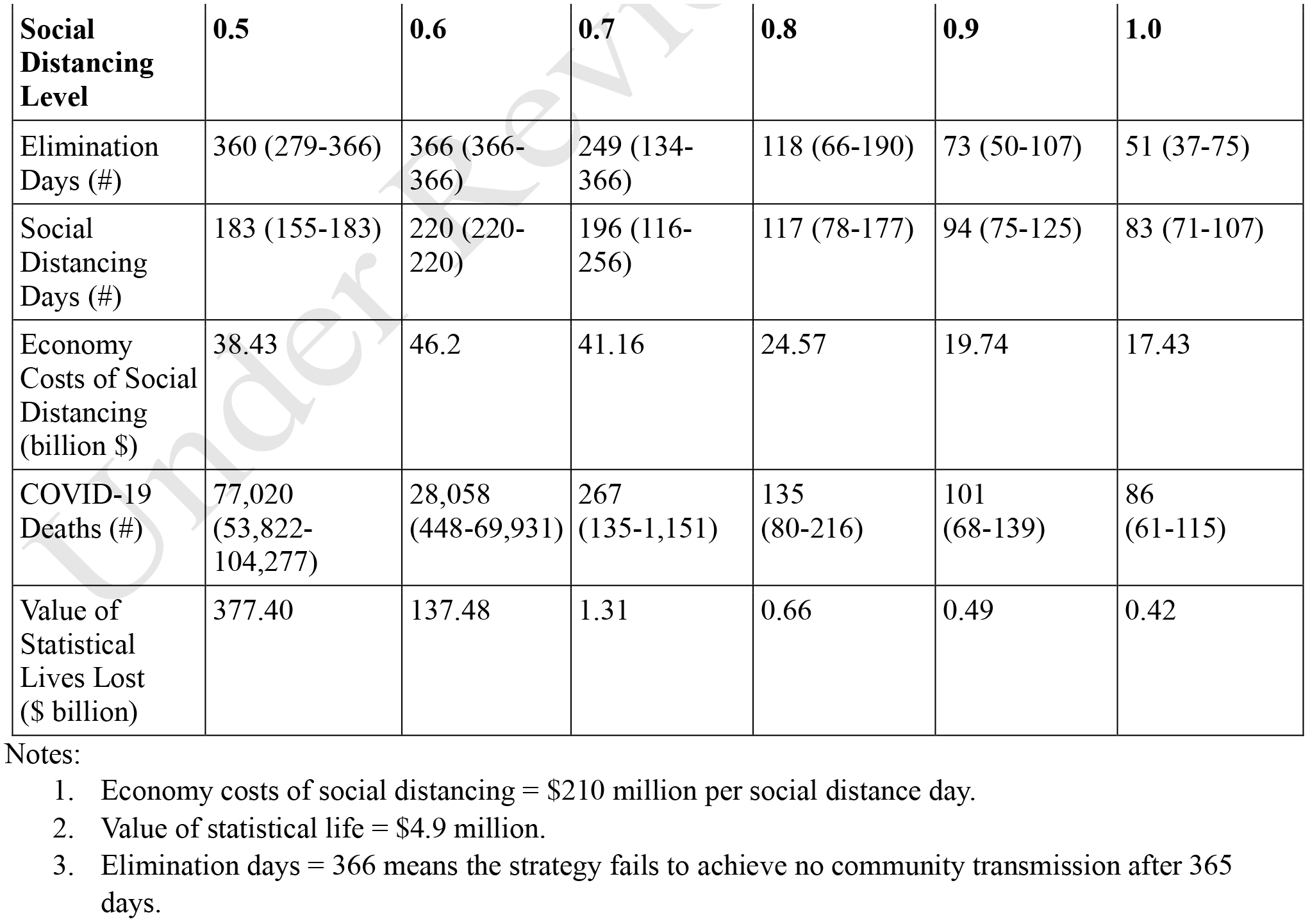
Median (2.5%-97.5 CI) values of additional Elimination Days and Social Distancing Days (sum of social distancing level each day for 365 days) and number of COVID-19 deaths and associated (based on median values) Economy Costs of Social Distancing, Value of Statistical Lives Lost and Hospitalisation Costs for Social Distancing Levels from 0.5 to 1.0 for 365 days after implementation of Social Distancing when average daily cases over the preceding 7 days exceeds 100.

| Social Distancing Level | 0.5 | 0.6 | 0.7 | 0.8 | 0.9 | 1.0 |
| --- | --- | --- | --- | --- | --- | --- |
| Elimination Days (#) | 360 (279-366) | 366 (366-366) | 249 (134-366) | 118 (66-190) | 73 (50-107) | 51 (37-75) |
| Social Distancing Days (#) | 183 (155-183) | 220 (220-220) | 196 (116-256) | 117 (78-177) | 94 (75-125) | 83 (71-107) |
| Economy Costs of Social Distancing (billion \$) | 38.43 | 46.2 | 41.16 | 24.57 | 19.74 | 17.43 |
| COVID-19 Deaths (#) | 77,020 (53,822-104,277) | 28,058 (448-69,931) | 267 (135-1,151) | 135 (80-216) | 101 (68-139) | 86 (61-115) |
| Value of Statistical Lives Lost (\$ billion) | 377.40 | 137.48 | 1.31 | 0.66 | 0.49 | 0.42 |

Notes:
1. Economy costs of social distancing = \$210 million per social distance day. 2. Value of statistical life = \$4.9 million. 3. Elimination days = 366 means the strategy fails to achieve no community transmission after 365 days.

On each day, the model steps through a sequence of processes, modifying the attributes of eligible cases. The attributes and processes have been designed to represent key features of COVID-19, and the effects of measures such as community testing, contact tracing and border controls. These processes are listed in Table 2 of the supplementary material, in the order they are applied within the daily time step. The IBM is fully stochastic at the individual level and the total number of infections produced by each infected individual is assumed to be a random variable drawn from a negative binomial distribution.

**Table 2.** Median (2.5%-97.5 CI) values of additional Social Distancing Days (sum of social distancing level each day for 365 days) and number of COVID-19 deaths and associated (based on median values) Economy Costs of Social Distancing, Value of Statistical Lives Lost and Hospitalisation Costs for Social Distancing Level = 1.0 for 365 days after implementation of Social Distancing.

|  | Relaxation Scenario A<sup>3</sup> | Relaxation Scenario B<sup>4</sup> |
| --- | --- | --- |
| Social Distancing Days (#) | 101 (71-210) | 115 (52-225) |
| Economy Costs of Social Distancing (billion \$) | 21.21 | 24.15 |
| COVID-19 Deaths (#) | 124 (66-261) | 190 (67-411) |
| Value of Statistical Lives Lost (\$ billion) | 0.61 | 0.93 |

Notes:
1. Economy costs of social distancing = \$210 million per social distance day. 2. Value of statistical life = \$4.9 million. 3. Social distancing is implemented for 40 days after which gradual relaxation over 60 days occurs once the weekly average of new daily recorded cases declines to 20. - No minimum of 40 days of social distancing; gradual relaxation over 60 days occurs once the weekly average of new daily recorded cases declines to 20.

#### Stochastic Compartment Model (SCM)

A parallel stochastic compartment model (SCM) replicates the behaviour of the IBM with high numbers of active cases. Instead of representing individuals, this model represents numbers of individuals in daily cohorts. Whereas the IBM assigns attributes to individuals, the SCM subdivides the total number of newly infected in each daily cohort into a set of compartments, detailed in Table 4 of the supplementary material. In the IBM, individuals can have different combinations of attributes, but the SCM must assign a unique compartment to each relevant combination of attributes. The SCM processes and corresponding exchanges between compartments are listed in Table 5 of the supplementary material.

#### Deterministic Compartment Model (DCM)

The DCM is identical in structure, parameters, and processes to the SCM. On every occasion where the SCM draws a random variable, the DCM replaces this with the expected value. It does not necessarily follow that case numbers predicted by the DCM equal the mean of case numbers in stochastic ensembles from the SCM, but in practice the differences between DCM trajectories and the median of SCM ensembles of trajectories are small.

#### Contact tracing

Contact tracing and enforced self-quarantine of downstream contacts assist in suppression at low case numbers. Their effectiveness is undermined by non-compliance of self-quarantine and lack of co-operation in terms of contact tracing that results in hidden transmission. Model fitting of parameters allow for results that account for both non-compliance and hidden transmission.

#### Model parameters

The IBM, SCM and DCM models share 22 model parameters that are provided in Table 6 of the supplementary material. A Bayesian inference procedure was used to fit the DCM to Australian observations obtained from https://www.covid19data.com.au/ and https://www.worldometers.info/coronavirus/#countries for the period February 20 to July 5, 2020. This procedure allowed us to obtain a posterior distribution for values of those parameters regarded as uncertain.

Model fitting allowed the data to inform the parameter values relating to hidden transmission. We find that hidden transmission (amplification of cases in populations with a low probability of presenting for testing) may have accounted for between 30 and 60% of new infections in the June-July resurgence in Victoria.

### Economy costs

Economy-wide costs of the national lockdown that began in March 2020 are based on Australian Bureau of Statistics (ABS) data at a Victorian level equivalent to approximately $210 million per lockdown day.^4^ Economy costs of a lockdown were assumed to be linear in the different levels of mandated social distancing, noting that greater social distancing and an increased frequency of lockdown (lockdown-relaxation-lockdown) increase economy costs. COVID-19 related fatalities are valued at $4.9 million per value of statistical life (VSL), sourced from Prime Minister and Cabinet^10^.

### Model simulations

Model simulations compared different mandated levels of social distancing assumed to be implemented when the average daily number of new recorded COVID-19 cases over the previous 7 days reached 100. Mandated social distancing measures were reintroduced on 9 July 2020 in Victoria, which coincides with the weekly average of new daily recorded cases of 100.

For each social distancing level, mandated measures remain in place for a minimum 40-day period and then social distancing is relaxed in a linear fashion over 60 days. Each level of social distancing (0.5, 0.6, 0.7, 0.8, 0.9 and 1.0) is benchmarked as a proportion of the estimated level of social distancing prevalent in Australia in April 2020 (see Table 1). All social distancing levels assume strong and enforced border controls for all new arrivals into Australia. In each scenario, social distancing is not relaxed until there is no recorded community transmission. Thus, each of the six cases in Table 1 assumes the goal is to achieve no community transmission.

Two relaxation scenarios, each with a social distancing level of 1.0, were also simulated (see Table 2). In each scenario, it is assumed social distancing measures are imposed when the weekly average of new daily recorded cases is 100 but relaxation may begin whenever the weekly average of new daily recorded cases declines to a trigger level of 20. In relaxation scenario A, the social distancing is implemented for a minimum of 40 days before the trigger level is assessed while in Scenario B there is no minimum period for social distancing. For both scenarios, once the relaxation criteria are met, gradual relaxation to zero social distancing occurs over a 60-day period. In each scenario, border quarantine leakage (failure) occurs at the probability of 0.2% per arrival from overseas (P_Q_ = 0.002).

## 3. Results

### Social distancing outcomes

A comparison of results for different levels of social distancing for 365 days after social distancing is first implemented are given in Table 1, assuming 0% leakage from hotel quarantine. The results are generated from the SCM with ensembles of 1,000 members drawn randomly from the Bayesian posterior distributions of the parameter set.

Elimination days is the number of days until *zero* community transmission (elimination) is achieved (value of 366 means elimination is *not* achieved) and social distance days is the sum of the level of social distancing on each day over 365 days. The scenarios with social distancing of 0.5 and 0.6 resulted in uncontrolled COVID-19 outbreaks. For social distancing of 0.5, some ensemble members achieve elimination within 365 days through herd immunity but at the loss of between 54,000 and 104,000 lives (Table 1).

Fig. 2a,b represents the simulations (median, quartiles, 5-95 percentiles) from a 1,000 ensemble and observed daily new local Australian cases for social distancing levels, respectively, 1.0 and 0.7. A social distancing level of 1.0 achieves no community transmission after about 50 days (median value) and within 80 days for *every* simulation. This level of social distancing results in economy costs of $17.4B. A social distancing level of 0.7 takes more than 250 days (median value) to achieve no community transmission and 21% of simulations fail to eliminate community transmission within one year, with costs estimated at $41.2B.

**Fig. 2a:**
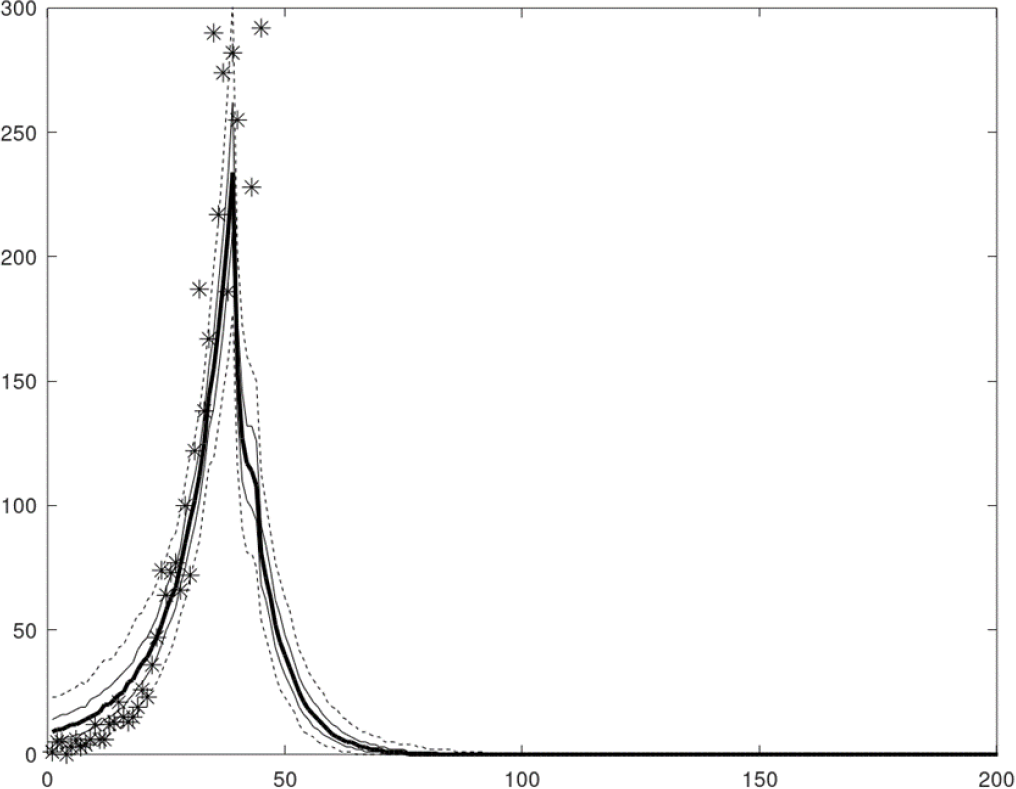
Elimination strategy, social distancing trigger = 100 daily cases, social distancing = 1.0 applied at day 35, and no leakage from border quarantine (P_Q_ = 0). Ensemble percentiles: median (thick line), quartiles (thin lines), 5-95 % percentiles (dashed lines), observed daily new Australian local cases, June 6 to 15 July 2020 (*).

**Fig. 2b:**
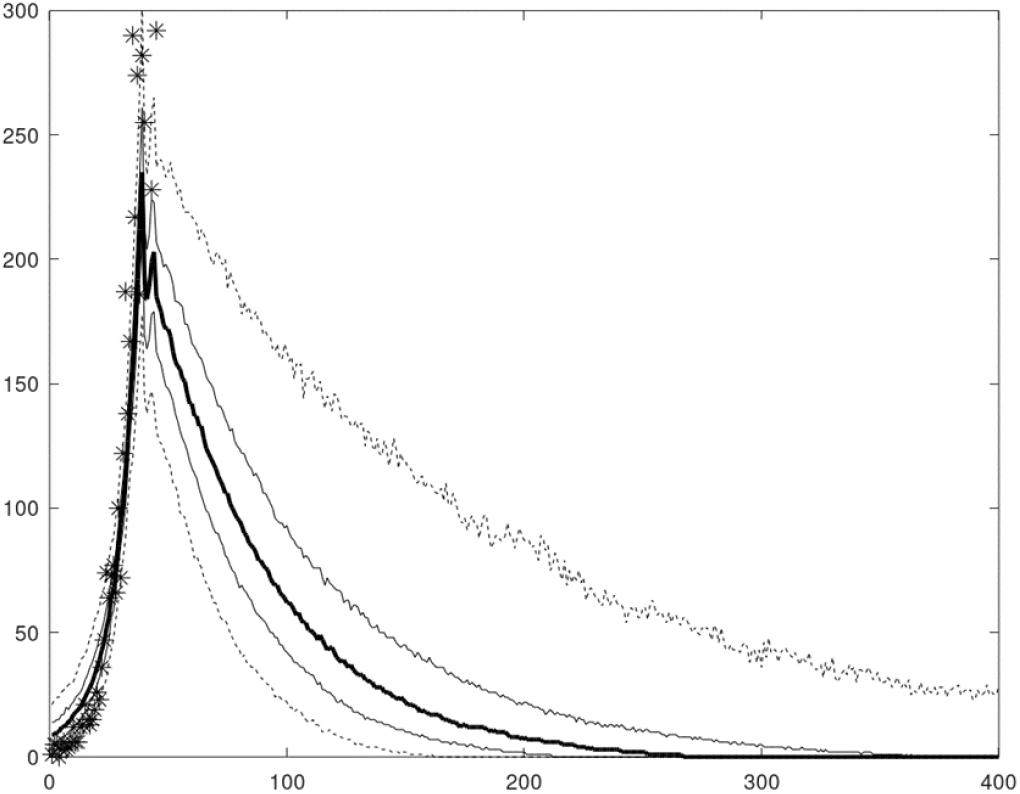
Elimination strategy, social distancing trigger = 100 daily cases, social distancing = 0.7 applied at day 35, and no leakage from border quarantine (P_Q_ = 0). Ensemble percentiles: median (thick line), quartiles (thin lines), 5-95 percentiles (dashed lines), observed daily new Australian local cases, June 6 to 15 July 2020 (*).

### Relaxation outcomes

Fig. 3a,b represents the simulations (median, quartiles, 5-95 percentiles) from a 1,000 ensemble and observed daily new cases for Suppression Scenario A and Scenario B. Both scenarios show a ‘yoyo’ pattern of outbreak-suppression-outbreak after the initial suppression with the 95 percentile simulations, but Scenario B generates substantially higher peaks in new daily cases. The median outcome achieves elimination within 100 days for Scenario A (Table 2), while it takes twice as long for Scenario B. Random border quarantine leakage contributes to multiple outbreaks, and especially for Scenario B (Fig. 3b) due to early relaxation and a failure to eliminate community transmission.

**Fig. 3A.**
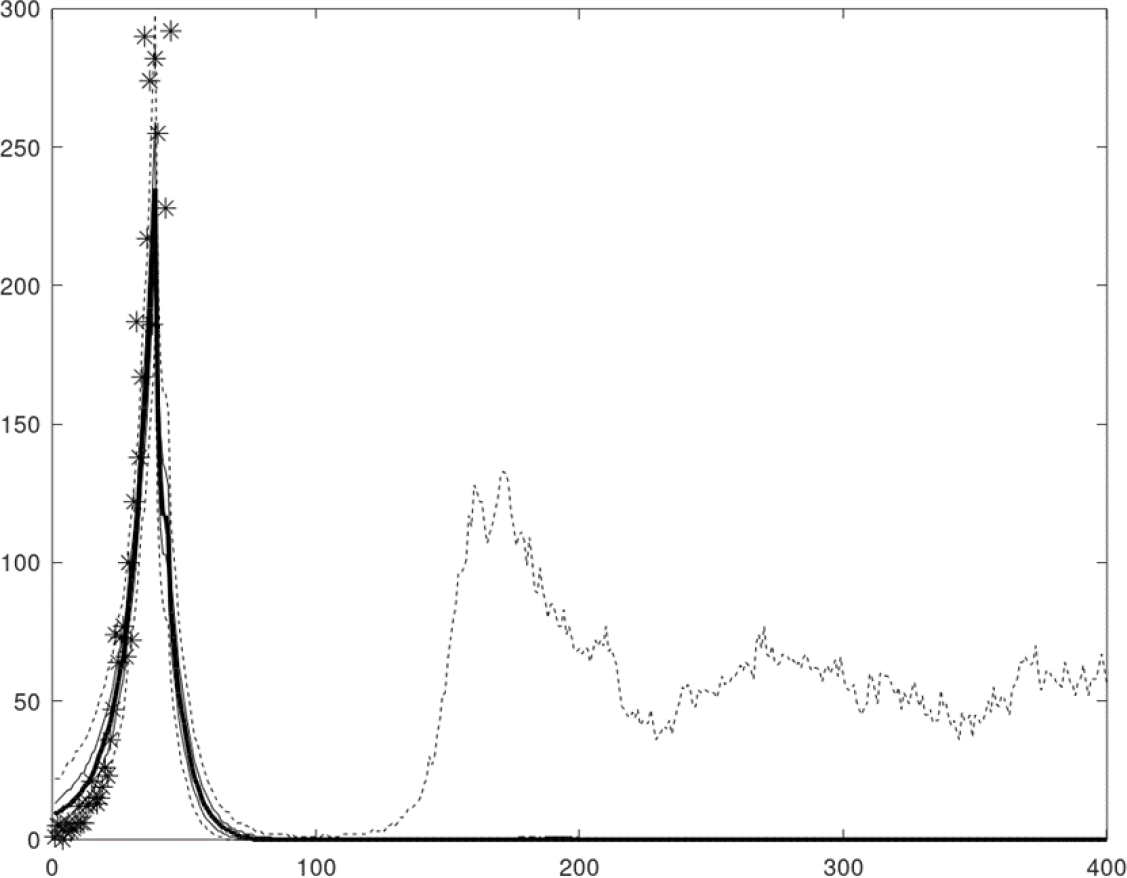
Suppression strategy, social distancing trigger = 100 daily cases, social distancing = 1.0 applied at day 35, minimum social distancing period = 40 days, relaxation trigger = 20 daily cases, and a very low probability of leakage from border quarantine (P_Q_ = 0.002). Ensemble percentiles: median (thick line), quartiles (thin lines), 5-95 percentiles (dashed lines), observed daily new Australian local cases, June 6 to 15 July 2020 (*).

**Fig. 3B.**
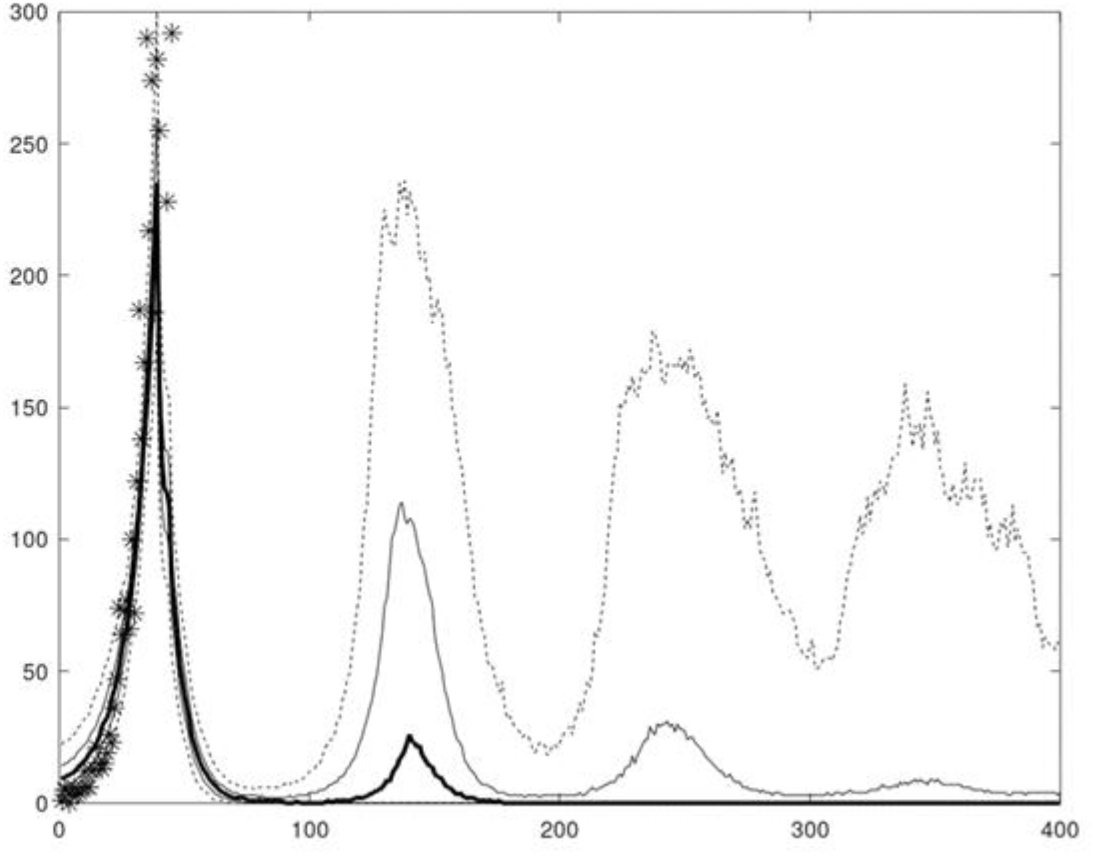
Suppression strategy, social distancing trigger = 100 daily cases, social distancing = 1.0 applied at day 35, relaxation trigger = 20 daily cases, no minimum social distancing period, and a very low probability of leakage from border quarantine (P_Q_ = 0.002). Ensemble percentiles: median (thick line), quartiles (thin lines), 5-95 percentiles (dashed lines), observed daily new Australian local cases, June 6 to 15 July 2020 (*).

## 4. Discussion

Our results provide robust support for a ‘Go Long and Go Hard’ strategy to optimise control COVID-19 infections in Australia. We find: one, that better public health outcomes (reduced COVID-19 fatalities) are *positively* associated with lower economy costs and higher levels of social distancing; two, achieving zero community transmission *lowers* both public health and economy costs compared to allowing community transmission to continue; three, early relaxation of social distancing, and in particular in the absence of a minimum social distancing period (minimum 40 days) and with quarantine leakage, *increases* both public health and economy costs.

We provide two sets of results. The first set in Table 1 assumes the goal of social distancing is achieving zero community transmission (elimination). We find that social distancing levels of 0.8, 0.9 and 1.0, as given in Table 1, achieve elimination with a 100% probability over the 365 days. Social distancing levels of 0.5 (in the absence of heard immunity) and 0.6 fail to achieve elimination within the simulation period. A social distancing level of 0.7 achieves elimination within 365 days with approximately 80% of simulations. We find that if reducing community transmission to zero is the goal, then lower levels of social distancing *increase* both COVID-19 fatalities and economy costs. This finding for Victoria is consistent with an agent-based model that compares a standard lockdown (with and without masks) with a more severe lockdown^11^.

The second set of results in Table 2 assumes that suppression is the goal such that relaxation of social distancing measures at 1.0 (assumed for both scenarios) does not begin until the weekly average of new daily recorded cases is 20, subject also to a minimum duration of 40 days for Scenario A. Table 2 results highlight the *lower* costs incurred when social distancing is imposed for a sufficiently long enough minimum period. Paradoxically, imposing a binding minimum number of social distancing days *reduces* the social distancing days over a 12-months period and, thus, the associated economy costs.

Our model suite does not allow us to fully capture the differences in transmission across different communities or sub-populations. Such transmission differences may arise from multiple factors including cultural reasons, housing density, and the proportion of workers who are in the casual workforce and who may have financial incentives not to be tested or go to work sick. The relationship between hidden transmission and essential workers is relevant for the effectiveness of social distancing because, depending on the severity measures (stage 3 versus stage 4), workers may still be able to infect their workmates at their workplaces.

## Data Availability

All relevant data is contained in the paper and the technical appendix.

## Technical Appendix

### A. Context

This technical appendix represents a summary of estimation and modelling undertaken over the period May to July 2020 by a team of researchers in response to a series of questions related to COVID-19 infections in Australia. The modelling was led by John Parslow with assistance from Kathryn Glass, Quentin Grafton and Tom Kompas, and advice from Emily Banks and Kamalini Lokuge.

This technical appendix provides a full description of the assumptions, parameters, data, and performance of the models by the research team. Results and simulations from the models will be included in two research papers currently under preparation.

### B. Conflict of Interest

None of the research team received any direct funding for this modelling and research. None of the research team declare any conflict of interest in relation to this work.

### C. Data

The data used for the estimation of model parameters come from https://www.covid19data.com.au/. Additional data (for early February-March data) come from the Worldometer website: https://www.worldometers.info/coronavirus/#countries.

### D. Model Description

The stochastic compartment model described here is one of a suite of epidemiological models developed to simulate the response of Covid-19 outbreaks to a range of control measures. These models are designed to offer a flexible and efficient representation of the progression of the disease in individual cases, transmission of the disease within the susceptible population, and the effects of community testing, downstream contact tracing, self-isolation and quarantine on detection and transmission. The stochastic models in particular are designed to provide insight into the effect of control measures on suppression and elimination at low case numbers.

The model suite comprises an individual-based model (IBM), a stochastic compartment model (SCM), and a deterministic compartment model (DCM). All three are numerical models that operate in discrete time with daily time steps.

#### D1. The Individual-based Model

The IBM follows infected individuals through time, starting on the day when they are infected, and ending when they are officially recovered. Each individual is characterised by a set of attributes whose values evolve over time according to a set of rules. All attributes, with one exception, take logical values of zero or 1, and determine the individual’s status with respect to source of infection (i.e. local or overseas), infectivity, display of symptoms, detection, traceability, self-isolation, self-quarantine or quarantine, recovery, hospitalization, and/or death. The attributes are listed in Table 1. The default values of all attributes are set to zero, until they are modified by model processes.

Only positive cases are represented in the model. All individual cases are represented in one long (NxM) array, where N is the number of cases and M the number of attributes. Cases are indexed in the array in order of creation, and new cases are added to the array as they occur through local transmission or arrival from overseas. The model also stores the first case index on each day t, IND1(t), and the number of cases on each day, NC(t), so that it can easily access all cases according to days since infection.

On each day, the model steps through a sequence of processes, modifying the attributes of eligible cases. The attributes and processes have been designed to represent key features of Covid-19, and the effects of measures such as community testing, contact tracing and border controls, as follows.

*Asymptomatic cases:* There is evidence that a proportion P_A_, somewhere between 10 and 40%, of infected individuals never display symptoms. The evidence suggests these individuals have lower viral loads and are less infectious than other cases. Individuals in the model are randomly assigned this attribute with probability P_A_ on the day they are infected. These individuals never display symptoms and are assigned a daily transmission coefficient G_A_ which is a fraction F_A_ of cases which develop symptoms.

*Timing of infectivity:* All newly infected individuals are non-infectious, and become infectious after T_I_ days, and then non-infectious after T_F_ days. The infectious attribute is switched on before transmission takes place on day T_I_, and off after transmission takes place on day T_F_, so individuals are infectious for T_L_ = T_F_+1-T_I_ days.

*Display of symptoms:* Individuals who are not asymptomatic display symptoms starting on day T_S_ and continue to display symptoms until they recover. Individuals displaying symptoms are potentially subject to testing and detection.

*Cases imported from overseas:* Detected overseas cases are introduced as “newly infected” cases T_S_ days prior to their detection and are ‘detected’ T_S_ days post-infection. This allows for the possibility of pre-symptomatic infection by overseas arrivals prior to the establishment of border controls and ensures daily detected overseas cases in the model match observations. Only symptomatic overseas cases are tested and detected. Thus, the model assumes reported overseas cases are matched by asymptomatic cases in the ratio P_A_:1-P_A_.

*Border controls:* From March 17, 2020, Australia required all overseas arrivals to self-quarantine at home, and from March 28, 2020, Australia required all overseas arrivals to undergo compulsory quarantine in hotels.

*Quarantine breakdown:* Cases in hotel quarantine are assumed not to contribute to transmission. To allow for a possible breakdown of hotel quarantine, quarantined individuals are converted into hidden community cases with a (very low) probability P_Q_.

*Community detection:* Individuals with Covid-19 symptoms can voluntarily report for testing and detection if positive. The model assumes cases with symptoms in the community are detected with daily probability P_DC_. Detected cases are required to self-isolate at home.

*Contact tracing:* The model records the array index QID of the infective source for each local case. If that source is detected, the downstream case is subject to contact tracing with daily probability P_T_. All cases identified as downstream contacts, including asymptomatic and pre-symptomatic cases, are required to self-quarantine at home.

*Contact tracing capacity:* The model sets a limit to contact tracing capacity, TCAP, measured in new (local) detected cases per day. Once daily local detected cases YDT exceed TCAP, the daily contact tracing probability is reduced by the factor TCAP/YDT.

*Detection in self-quarantine:* Once cases in self-quarantine display symptoms, they are assumed to be tested and detected with (high) daily probability P_DSQ_. Detected cases are required to self-isolate at home.

*Hospitalization:* Symptomatic cases develop complications requiring hospitalisation on day T_H_, with probability P_H_. Hospitalized individuals remain in hospital until they recover or die.

*Fatally ill and deceased:* On day T_D_, hospitalised patents are assigned the fatally ill attribute with probability P_M_. From day T_D_ onwards, fatally ill patients die with daily probability P_D_. This allows a long exponential ‘tail’ of deaths, which matches observations.

*Recovery:* From day T_R_ onwards, all cases that are not fatally ill or deceased recover with daily probability P_R_. Again, this allows a long exponential tail of recoveries. Because transmission ends T_F_ days after infection, recovery in the model serves primarily as a reporting mechanism.

*Hidden or uncooperative cases:* Contact tracing can be extremely effective in amplifying even low rates of community detection and shutting down transmission. Observations suggest it may be less effective in practice. The model allows for a fraction P_U_ of hidden or uncooperative individuals who evade both community detection and downstream contact tracing, to allow more realistic levels of contact tracing effectiveness.

These processes are listed in Table 2, in the order they are applied within the daily time step. Cases are considered active until the number of days post-infection (d) exceeds the maximum time to recovery T_M_. In practice, only cases having eligible values of d are considered in applying each process.

The IBM is fully stochastic at the individual level, so whenever a process imposes a change with probability P, that change occurs for an individual if and only if U < P, where the random variable U ~ Uniform(0,1).

##### Transmission

This IBM is similar to other individual-based epidemiological models in its representation of the evolution of the disease. Large and computationally expensive individual-based models structure the whole population according to a predefined social network of contacts and potential transmissions. Our model employs a simpler representation of transmission, in which new infections are assumed to come from a large, well-mixed susceptible population and are added to the indexed array of positive cases. This allows this model to represent only positive cases and incur low computational costs, at least for small outbreaks.

The total number of infections produced by each infected individual is assumed to be a random variable drawn from a negative binomial distribution. The negative binomial is widely used to represent the distribution of lifetime infections and daily contacts. The distribution is characterised by the mean (reproductive number) R, the dispersion coefficient k, and the derived probability P_B_ = 1 - k/(R+k). For each case, the total number of people they infect is the sum of infections on each of T_L_ successive days, where T_L_ is the infectious period. The sum of n random variables drawn from a negative binomial with parameters p and k’ is itself a negative binomial with parameters p and n.k’. Thus, the daily infections for each individual are drawn from a negative binomial with parameters P_B_ and k’= k/T_L_. The daily mean transmission rate G = R/T_L_, so

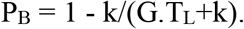

The distribution of lifetime infections is believed to be over-dispersed, and so the distribution of daily infections is then highly over-dispersed. Most infected individuals produce zero infections on any given day, while ‘super-spreader’ individuals can produce large clusters of 30 or 50 infections in one event with non-negligible probability.

The daily mean transmission rate G is multiplied by the ratio of the remaining susceptible population S(t) to the total population, POP. In the simulations considered in this study, S/POP remains very close to 1. The mean transmission rate also depends on citizen behaviours and can be reduced by a variety of social distancing measures, discussed below. For asymptomatic cases, G is reduced by the multiplier F_A_. The parameter P_B_ is adjusted to match G and k.

Quarantined cases in hotels are assumed not to contribute to transmission (unless a rare quarantine breakdown converts a quarantined case to a hidden case.) Violations of self-quarantine and self-isolation at home are assumed to be more common, and these cases are assumed to contribute to daily transmission with (low) probability P_L_.

The transmission module considers all cases which satisfy T_I_<=d<=T_F_. It rejects cases with QQ = 1or QI=0. For accepted cases with QA = 0, it draws the number of daily infections from the negative binomial with parameters G and k, as described above. For accepted cases with QA = 1, it draws the number of daily infections from the negative binomial with parameters G.F_A_ and k.

##### Outputs

As it carries out the processes in Table 2, the IBM records a set of daily output variables listed in Table 3.

#### D2. The Stochastic Compartment Model

While the IBM has been designed to be computationally efficient, the computational cost of processing individual cases is still large when uncontrolled epidemics lead to millions of active cases. A parallel stochastic compartment model (SCM) has been developed to replicate the behaviour of the IBM efficiently with high numbers of active cases. Instead of representing individuals, this model represents numbers of individuals in daily cohorts. On day t, the SCM represents the number of individuals that were newly infected on day t_0_ as X(t_0_,t), for values of d = t – t_0_ (days post-infection) from 0 to the maximum recovery time, T_M_.

Whereas the IBM assigns attributes to individuals, the SCM subdivides the total number X(t_0_,t) in each daily cohort into a set of compartments, X_i_(t_0_,t). In the IBM, individuals can have different combinations of attributes, but the SCM must assign a unique compartment to each relevant combination of attributes. Consequently, the number of SCM compartments needs to be larger than the number of IBM attributes. The compartment labels and descriptions are given in Table 4. The changes in individual attributes with days post-infection in the IBM are represented in the SCM by the movement of appropriate numbers of individuals between compartments. When changes in individual attributes in the IBM occur randomly with probability P, the number of individuals transferred between corresponding compartments in the SCM is drawn from a binomial distribution with parameters N and P, denoted here by BIN(N,P), where N is the number potentially eligible for transfer. This means that the SCM exactly replicates the behaviour of the IBM in a statistical sense for all processes except for transmission and contact tracing.

The SCM processes and corresponding exchanges between compartments are listed in Table 5, again in the order of execution in daily time steps. Although the notation is different, the SCM processes for the most part have identical effect to those described for the IBM, and the description and justification provided earlier applies equally here.

##### Transmission

The number of potentially infectious sources on day t is obtained by summing cases from all potentially infectious cohorts X(t_0_,t) with T_I_ <= t – t_0_ <= T_F_. These sources need to be divided into five classes, according to their contributions to transmission:

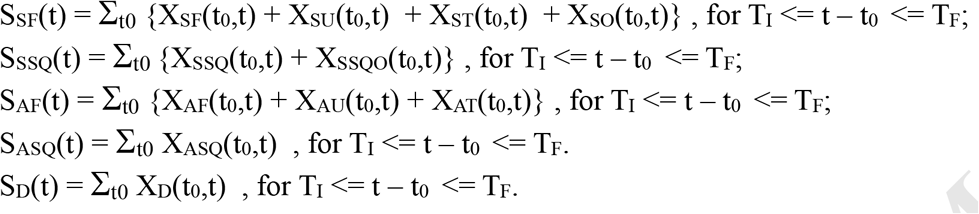

These sources are then weighted appropriately to calculate their expected contribution to transmission. Contributions from cases in self-isolation and self-quarantine at home are multiplied by the proportion P_L_ that breach self-isolation / self-quarantine. Contributions from asymptomatic community cases are multiplied by F_A_. These scaled contributions are then summed to produce an infectious potential on day t, IP(t):

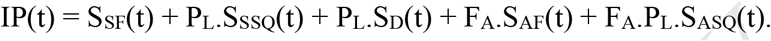

Daily infections per individual are still assumed to have a negative binomial distribution with parameters P_B_ and k’= k/T_L_. The sum of daily infections from IP(t) sources will then be negative binomial with parameters P_B_ and k’.IP(t). This produces a probability distribution for the number of new local infections which is similar to that for the IBM. (The IBM computes the contributions from isolated individuals as a random variable with expected value P_L_.X_SI_.)

##### Contact Tracing

The SCM also differs from the IBM in its representation of downstream contact tracing. In the IBM, the index of the infection source for each active case is known, and it is a simple matter to assess whether the source has been detected. In the SCM, only the proportion of detected cases among potential sources for any given cohort is known. Thus, the fraction of detected potential sources cannot be used directly as the fraction of the cohort subject to downstream contact tracing. The quantity IP(t) used above to calculate transmission is the appropriate weighted sum of infectious potential over source cohorts for the cohort formed on day t.

A cohort X(t_0_,t) on day t has potential source cohorts X(t_1_,t) with T_I_ <= t_0_ – t_1_ <= T_F_. We calculate the proportion of the source potential IP(t_0_) which is represented by the detected components X_D_(t_1_,t) among these relevant source cohorts on day t. Under the model assumptions, no asymptomatic cases are detected, and hotel quarantined cases produce zero infections. Thus, we need only separate the detected sources X_D_(t_1_,t) into those that were free or ‘traceable’ on day t_0_, D_F_(t_0_,t), and those that were self-quarantining on day t, D_SQ_(t_0_,t). This can be done as follows.

In processing transmission on each day t_0_, the model computes and stores the total number of self-quarantining symptomatic sources S_SSQ_(t_0_), and the total number of detected sources S_D_(t_0_), summed over potential source cohorts. On day t = t_0_ + d, the same calculation can be done over the updated source cohorts X(t_1_,t) of cohort X(t_0_,t), to produce S_SSQ_(t_0_,d), S_D_(t_0_,d). Because self-isolated symptomatic cases are detected with high probability, it is reasonable to assume that the increase in detected sources comes first from the self-isolated symptomatic source pool on day t_0_, S_SSQ_(t_0_), and after that from the free or traceable source pool S_SF_(t_0_). So:

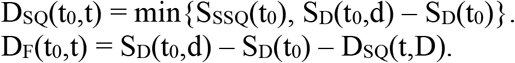

The reduction in infectious potential since t_0_ is then:

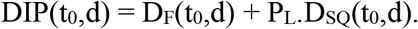

The fraction of cohort X(t_0_,t) subject to downstream contact tracing on day t should then be:

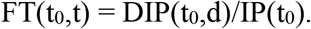

Individuals are moved incrementally from free to traceable compartments in each daily update. The fraction moved each day should be proportional to the daily increase in the traceable fraction, FT(t_0_,d) – FT(t_0_,d-1), with FT(t_0_,0) = 0. The number to be moved is calculated as a proportion of the number remaining. Thus, the fraction moved each day from AF to AT and SF to ST is:

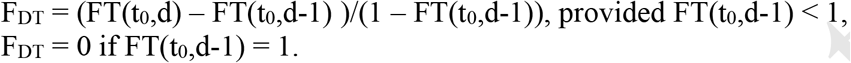

The numbers moved each day are DXST = F_DT_.XSF(t_0_,t), DXAT = F_DT_.XAF(t_0_,t) (The SCM deals in integer numbers of cases, so DXST, DXAT are rounded to the nearest integer. If either is less than 1, it is set to 1 with probability DXST or DXAT, zero otherwise).

#### D.3 The Deterministic Compartment Model (DCM)

The DCM is identical in structure, parameters and processes to the SCM. On every occasion where the SCM draws a random variable from the binomial or negative binomial, the DCM replaces this with the expected value. For the binomial, this means replacing Z ~ BIN(N,P) by N.P. It does not necessarily follow that case numbers predicted by the DCM will equal the mean of case numbers in stochastic ensembles from the SCM, but in practice the differences between DCM trajectories and the median of SCM ensembles of trajectories are observed to be small.

The ‘number’ of cases in compartments in the DCM are real numbers rather than integers, so it cannot reproduce random small number effects or predict probabilities of elimination. The DCM is more than an order of magnitude faster computationally than the SCM and emulates the SCM quickly and usefully in many situations. It has proved particularly useful as a fast emulator for Bayesian inference, as discussed in the section ‘Model Parameters’.

**Table 1.** Individual Attributes in IBM

| Attribute | Name | Significance |
| --- | --- | --- |
| QA | Asymptomatic | 1 if asymptomatic, 0 otherwise. |
| QI | Infectious | 1 if infectious, 0 otherwise. |
| QS | Symptomatic | 1 if symptomatic, 0 otherwise |
| QH | Hospitalized | 1 for severe hospitalized cases, 0 otherwise |
| QF | Fatally ill | 1 if fatally ill, 0 otherwise |
| QM | Dead | 1 if dead, 0 otherwise |
| QR | Recovered | 1 if recovered, 0 otherwise |
| QO | Overseas Origin | 1 if arrived from overseas, 0 if local |
| QQ | Quarantine | 1 if placed in hotel quarantine, 0 otherwise |
| QSQ | Self-isolating or self-quarantined | 1 if instructed to self-isolate or self-quarantine at home, 0 otherwise |
| QD | Detected | 1 if tested positive, 0 otherwise |
| QID | Source index | Array index of the infective source |
| QU | Hidden / unco-operative | 1 if hidden from community testing and contact tracing, 0 otherwise. |

**Table 2.** Processes modifying attributes in IBM, in order of execution in each daily time step.

| Process | Daily cohorts | Eligible cases: Effect, Probability |
| --- | --- | --- |
| Start infectious | $d = T_I$ | All: $QI = 1$ |
| Stop infectious | $d = T_F + 1$ | All: $QI = 0$ |
| Transmission | $T_I \leq d \leq T_F$ | $QI = 1$: See Text. |
| Source index | $d = 0$ | New local cases: $QID =$ array index of infection source. |
| Asymptomatic | $d = 0$ | New local cases: $QA = 1$ with probability $P_A$ |
| Hidden | $d = 0$ | New local cases: $QU = 1$ with probability $P_U$ |
| Overseas arrival | $d = 0$ | Detected overseas arrivals: $QO = 1$. (Overseas cases are introduced as new infections, $T_S$ days before detection.) |
| Overseas arrivals, asymptotic | $d = 0$ | For each detected overseas arrival, an asymptotic arrival is created with probability $P_A/(1-P_A)$: $QA = 1$, $QO = 1$. |
| Hotel quarantine | $d = 0$ | All overseas arrivals after March 28: $QQ = 1$. |
| Hotel-quarantine breakdown | $d = 0$ | Cases in quarantine, $QQ=1$: $QQ = 0$, $QH = 1$ with probability $P_Q$. |
| Overseas self-quarantine | $d = 0$ | Overseas arrivals from March 17 to 28: $QSQ = 1$. |
| Contact tracing and self-quarantine | $d \geq 1$ | Local non-hidden cases ($QU = 0$) with detected sources, $QD(QID)=1$: $QSQ = 1$, with daily probability $P_T$. |
| Detection of overseas cases | $d = T_S$ | Overseas arrivals, $QO = 1$, $QA = 0$: $QD = 1$, $QSQ = 1$. |
| Detection of cases in self-quarantine. | $d \geq T_S$ | For $QSQ = 1$, $QA = 0$: $QD = 1$ with daily probability $P_{DSQ}$ |
| Detection of cases in community | $d \geq T_S$ | For $QA = 0$, $QU = 0$, $QSQ = 0$: $QD = 1$, $QSQ = 1$ with daily probability $P_{DC}$ |
| Hospitalized | $d = T_H$ | For $QA = 0$: $QH = 1$, $QD = 1$ with probability $P_H$ |
| Fatally ill | $d = T_D$ | For $QH = 1$: $QF = 1$ with probability $P_M$ |
| Death | $T_D \leq d \leq T_M$ | For $QF = 1$: $QM = 1$ with daily probability $P_D$ |
| Recovery | $T_C \leq d \leq T_M$ | For $QF = 0$, $QM = 0$: $QR = 1$ with daily probability $P_R$ |

**Table 3.** Output variables from IBM, SCM

| **Variable name** | **Description** |
| --- | --- |
| Y0 | Daily new cases created(total) |
| Y0L | Daily new cases created (local) |
| YDT | Daily new detected cases |
| YDTL | Daily new detected cases (local) |
| YH | Hospital admissions |
| YHA | Active hospital cases |
| YM | Daily deaths |
| YRD | Daily detected recoveries. |
| Y0C | Cumulative cases |
| YDTC | Cumulative detected cases |
| YHC | Cumulative hospital admissions |
| YMC | Cumulative deaths |
| YRDC | Cumulative detected recoveries. |

**Table 4.** Compartments comprising daily cohorts in SCM

| **Label** | **Description** |
| --- | --- |
| SF | Symptomatic / pre-symptomatic in community |
| AF | Asymptomatic in community |
| SU | Hidden symptomatic / pre-symptomatic |
| AU | Hidden asymptomatic |
| ST | Traceable symptomatic / pre-symptomatic in community |
| AT | Traceable asymptomatic in community |
| SSQ | Symptomatic / pre-symptomatic in self-quarantine |
| ASQ | Asymptomatic in self-quarantine |
| D | Detected (symptomatic) cases |
| SFO | Overseas pre-symptomatic in community |
| SSQO | Overseas pre-symptomatic in self-quarantine |
| SQ | Overseas symptomatic / pre-symptomatic in hotel quarantine |
| AQ | Overseas asymptomatic in hotel quarantine |
| SH | Hospital patients |
| FI | Fatally ill hospital patients |
| M | Dead |
| RD | Recovering detected cases |

**Table 5.** Processes controlling compartment exchanges in SCM, in order of execution in each daily time step.

| Process | Daily Cohorts | Compartment exchanges |
| --- | --- | --- |
| Transmission | $T_I \leq d \leq T_F$ | See Text. |
| Asymptomatic & hidden | $d = 0$ | Given $X$ new local cases: $[X_{SF}, X_{AF}, X_{SU}, X_{AU}]$ is ∼ multinomial ($X$, $[(1-P_A).(1-P_U), P_A.(1-P_U), P_U(1-P_A), P_A.P_U]$) |
| Overseas arrivals | $d = 0$ | Overseas reported (symptomatic) cases $X_{SO}$ treated as newly infected $T_S$ days earlier. Matched by asymptomatic cases $X_{AO} \sim \text{BIN}(X_{SO}, P_A/(1-P_A))$. For arrivals:<br>Before March 17: $X_{SFO} = X_{SO}$, $X_{AF} = X_{AF} + X_{AO}$.<br>From March 17 to March 28: $X_{SSQO} = X_{SO}$, $X_{ASQ} = X_{AO}$.<br>After March 28: $X_{SQ} = X_{SO}$, $X_{AQ} = X_{AO}$. |
| Hotel quarantine breakdown | $d = 0$ | Hotel quarantine evasions $QE \sim \text{BIN}(X_{SQ}, P_Q)$.<br>$X_{SQ} = X_{SQ} - QE$, $X_{SU} = X_{SU} + QE$. |
| Source detected | $d \geq 1$ | $DX_{ST}$ = additional symptomatic cases with detected sources. $DX_{AT}$ = additional asymptomatic cases with detected sources. (See text for derivation).<br>$X_{ST} = X_{ST} + DX_{ST}$, $X_{SF} = X_{SF} - DX_{ST}$.<br>$X_{AT} = X_{AT} + DX_{AT}$, $X_{AF} = X_{AF} - DX_{AT}$. |
| Contact tracing | $d \geq 1$ | $CST \sim \text{BIN}(X_{ST}, P_T)$. $X_{ST} = X_{ST} - CST$, $X_{SSQ} = X_{SSQ} + CST$.<br>$CAT \sim \text{BIN}(X_{AT}, P_T)$. $X_{AT} = X_{AT} - CAT$, $X_{ASQ} = X_{ASQ} + CAT$. |
| Detection of overseas cases | $d = T_S$ | $X_D = X_D + X_{SFO} + X_{SSQO}$, $X_{SFO} = 0$, $X_{SSQO} = 0$.<br>$Y_{DET} = Y_{DET} + X_{SFO} + X_{SSQO} + X_{SQO}$ |
| Detection of cases in self-quarantine | $d \geq T_S$ | $DSQ \sim \text{BIN}(X_{SSQ}, P_{DSQ})$. $X_{SSQ} = X_{SSQ} - DSQ$, $X_D = X_D + DSQ$.<br>$Y_{DET} = Y_{DET} + DSQ$, $Y_{DETL} = Y_{DETL} + DSQ$. |
| Detection of cases in community | $d \geq T_S$ | $DSF \sim \text{BIN}(X_{SF}, P_{DC})$. $X_{SF} = X_{SF} - DSF$, $X_D = X_D + DSF$.<br>$DST \sim \text{BIN}(X_{ST}, P_{DC})$. $X_{ST} = X_{ST} - DST$, $X_D = X_D + DST$.<br>$Y_{DET} = Y_{DET} + DSF + DST$, $Y_{DETL} = Y_{DETL} + DSF + DST$. |
| Hospitalization | $d = T_H$ | For each of $X = X_{SF}, X_{SSQ}, X_{ST}, X_{SU}, X_D, X_{SQ}$<br>$DH \sim \text{BIN}(X, P_H)$. $X_{SH} = X_{SH} + DH$, $Y_H = Y_H + DH$.<br>For $CC = SF, SSQ, ST, SU, D$: $X_{CC} = X_{CC} - DH$.<br>For $CC = SF, SSQ, ST, SU$: $Y_{DET} = Y_{DET} + DH$, $Y_{DETL} = Y_{DETL} + DH$. |
| Fatally ill | $d = T_D$ | $DFI \sim \text{BIN}(X_{SH}, P_M)$. $X_{SH} = X_{SH} - DFI$, $X_{FI} = X_{FI} + DFI$. |
| Death | $T_D \leq d \leq T_M$ | $DD \sim \text{BIN}(X_{FI}, P_D)$. $X_{FI} = X_{FI} - DD$, $X_M = X_M + DD$.<br>$Y_M = Y_M + DD$. |
| Recovery | $T_C \leq d \leq T_M$ | $DRD \sim \text{BIN}(X_D, P_R)$. $X_D = X_D - DRD$, $X_{RD} = X_{RD} + DRD$.<br>$DRH \sim \text{BIN}(X_{SH}, P_R)$. $X_{SH} = X_{SH} - DRH$, $X_{RD} = X_{RD} + DRH$.<br>$Y_{RD} = Y_{RD} + DRD + DRH$. |

#### E. Model Parameters

The models described in the preceding section share 22 model parameters (Table 6). These can be divided into 4 categories.

The first group, T_S_, T_I_, T_F_, T_H_, T_D_, T_M_, specify the timing of the progression of the disease through infected cases, in days post-infection. This progression is now well studied for Covid-19, and there is broad agreement on typical or mean values of these parameters. Although it can take considerably longer, the typical time T_S_ to onset of symptoms is about 5 days, and the common assumption is most cases are infectious for approximately two days before onset of symptoms. We set T_I_ = 4 with T_S_ = 5 which ensures two rounds of transmission prior to onset of symptoms and possible detection. The duration of infection is less tightly defined but estimates of the Reproductive Number (R) are around 2.5, and uncontrolled epidemic growth rates can be as high as a doubling every 3 to 4 days. Setting T_F_ to 8 days allows the model to achieve these growth rates with R around 2.5.

Severe complications typically develop around 5 to 7 days after onset of symptoms, and T_H_ is set here to 10 days, T_D_ is set to 12 days and T_R_ to 19 days. These parameters were previously estimated in fitting a related compartment model to Australian data through to the end of April. It will be shown below that these values of T_D_ and T_R_ offer a reasonable fit to observations of cumulative deaths and recoveries in Australia. In practice, these three parameters have negligible effect on rates of transmission and the trajectory of daily cases, though they clearly have important implications for the hospital system. Similarly, the maximum duration T_M_ of 40 days is imposed for convenience and computational efficiency and has negligible effect on model outputs. Given the values adopted for other parameters, almost all cases are resolved before reaching 40 days post-infection.

The second group of parameters, P_A_, P_H_, P_M_, P_D_, P_R_, also control the nature and progression of the disease in individuals. There have been widely varying estimates of the proportion of cases which are asymptomatic, but it is now generally agreed that this fraction is less than 50%, and likely in the range 10 to 40%. In Australia’s ‘first wave’, the proportion of detected cases that were hospitalised was around 10%, and the overall proportion of deaths among hospital cases has been around 11%, so P_H_ is set to 0.1, and P_M_ to 0.11. The daily death and recovery rates P_D_ and P_R_ determine the time constants associated with the exponential tail of deaths and recoveries after T_D_ and T_R_ respectively. P_D_ is set to 0.15, and P_R_ to 0.2, corresponding to time constants of 6.7 and 5 days respectively. These provide reasonable agreement with observations (see Fig. 5, 6 below).

The mean daily transmission rate G0 in the absence of social distancing is a critical parameter, controlling the evolution of the initial outbreak. Values for this parameter, along with the relative transmission for asymptotic cases F_A_, are determined by fitting the deterministic model to Australian observations using Bayesian inference methods. A prior range for G0, from 0.3 to 0.65 is used there. When multiplied by TL = 5 days, this corresponds to Reproductive Numbers (R0) from 1.5 to 3.25, which includes most reported values. It has been argued that asymptomatic cases, including children, have transmission rates that are less than 50% of transmission rates in cases that go on to develop symptoms, and a prior range for FA of 0.1 to 0.4 is adopted here.

The next group of parameters, P_DC_, P_DSQ_, P_T_, P_L_, P_U_, P_Q_ and TCAP, determine the effectiveness of community testing, contact tracing, self-isolation, self-quarantine and hotel quarantine. These can be seen as control parameters, in the sense that they depend on the effort and efficiency of these measures, but they also depend on public behaviour and compliance. Here, we assume that the testing and detection of symptomatic cases already in self-quarantine is efficient and fast, so P_DSQ_ is set to a daily value of 0.8. The probability of hotel quarantine breakdown P_Q_ is treated as a parameter to be varied in scenarios, but is assumed to be very low, 0.01 or less. The probability of violation of self-quarantine and self-isolation leading to transmission, P_L_, is also low. There were early reports it could be around 20% and it is given a prior range of 0.1 to 0.3.

Community testing rates were low until late April in Australia, owing to a shortage of testing kits. Since May, community testing rates have been relatively high, and community members experiencing symptoms have been strongly encouraged to come forward for testing. Community detection rates P_D_ in this period are given a prior range of 0.2 to 0.5 per day. Contact tracing has been professionally trained and supported in Australia and, provided contact tracing capacity is not exceeded, one would expect downstream contact tracing to be relatively efficient. The proportion of downstream contacts traced per day, P_T_, is assigned a wide prior range of 0.2 to 0.8. The extent of public cooperation in voluntarily submitting for testing, and assisting contact tracing, is unknown, but recent events in Victoria have suggested uncooperative behaviour could be more widespread than we might hope. The fraction of hidden / uncooperative cases is assigned a wide prior range of 0.1 to 0.6.

The model measures tracing capacity in local detected cases per day. For each detected case, a number of downstream contacts of order 10 must be identified, contacted, asked to self-quarantine, and monitored for development of symptoms, and to check compliance, for 14 days. Thus, for 100 daily detected cases, contact tracers could have of order 14,000 contacts under management. It seems likely that the tracing capacity TCAP in Australia is between 100 and 500 daily detected cases (and would be less in each state or territory). For the data-fitting exercise described below, TCAP is set to 500, so tracing capacity is not limiting. It is set to lower values in some scenarios.

Finally, the susceptible population POP is set to 20 million for Australia, assuming approximately 80% of the population is initially susceptible.

##### Bayesian Inference

A simple Bayesian inference procedure was used to fit the DCM to Australian observations for the period February 20 to July 5, 2020 to obtain a posterior distribution for values of those parameters regarded as uncertain. Among the parameters discussed above, prior ranges were specified for G0, F_A_, P_A_, P_L_, P_T_, P_U_ and P_DC_.

In addition to the border control measures already incorporated in the model, Australia imposed strong social distancing measures, commencing March 16, 2020 and strengthened on March 23, 2020 to preclude all but essential movement outside the household. This will have reduced social contacts and the transmission rate G. It is represented in the model by a linear decline in G over a 2-week period starting March 16, 2020. The absolute reduction in G is uncertain, so the minimum value of G at the height of the lockdown is treated as another uncertain parameter G_LD_, with a prior range 0.05 to 0.25.

The lockdown was officially relaxed starting May 8, 2020, but Google and Apple movement data show a ‘spontaneous’ increase in movement starting in early April and continuing through the official relaxation period in more or less linear fashion. The model, therefore, assumes a linear increase in G from its minimum value G_LD_ to a final relaxed value G_R_ on July 5, 2020. However, if G_R_ is added as another parameter, the inference procedure yields very high positive correlations between G0 and G_R_. Instead, the model estimates the relative relaxation of social distancing by July 5, 2020 measured as RSD = (G_R_-G_LD_)/(G0-G_LD_). The parameter RSD is assumed to be less than 1 and given a prior range of 0.7 to 1.0.

Initial attempts at Bayesian inference yielded high positive posterior correlations between G0 and P_A_, when G0 was interpreted as the transmission rate for symptomatic cases. This is unsurprising, as the average transmission rate across symptomatic and asymptomatic cases is reduced when P_A_ increases. The estimated transmission rates G0 and G_LD_ are, therefore, defined as average rates across both symptomatic and asymptomatic cases. The initial transmission rate for symptomatic cases is given by G0/(1-P_A_.(1-F_A_)), and the rate for asymptomatic cases by F_A_.G0/(1-P_A_.(1-F_A_)). This transformation virtually eliminates posterior correlations between G0 and P_A_ or F_A_.

As discussed earlier, community testing rates have also changed over time. Community testing rates were low up until late April 2020 because of a shortage of testing kits, and because priority was given to overseas travellers and their contacts. Starting April 22, 2020 community testing rates increased markedly over a 2-week period. In fitting the observations, the model assumes community detection rates were low, equal to 0.2, prior to 22 April, 2020 and then increased over a 2-week period to a value P_DC_, to be estimated from a prior range of 0.2 to 0.5.

In fitting the observations, the model is driven by reported overseas cases, in the manner described in the preceding section. The model was assessed against local detected cases (i.e. reported cases minus overseas cases). Overseas cases represented about 70% of the March-April 2020 peak, and the model reproduces the input stream of overseas cases as detected cases, so fitting total cases could have been a poor test of the model.

A simple SIR (Sample Importance Resample) procedure was used to obtain the posterior distribution of parameters. The DCM was run using 200,000 independent random samples from the prior parameter distribution (the prior was treated as uniform on the parameter ranges specified in Table 6, and parameters were treated as independent in the prior). For each model run, a likelihood was calculated based on the SSQ errors between predictions and observations. To reduce any weekly reporting artefacts, and to allow for the DCM’s inability to reproduce any stochastic variation in the observations, time series of observations and predictions were smoothed by a running 7-day average before calculating the SSQ error. A ln(X+10) transform was also applied to the smoothed predictions and observations before computing the SSQ residuals, to give equal weight to errors at low and high case numbers, and to render the residuals approximately Gaussian. The likelihood was calculated from the SSQ assuming a Gaussian distribution of errors, with the degrees of freedom equal to the number of observations minus the number of parameters all divided by 7 to account for the effects of the 7-day running average. The error variance was estimated from the minimum SSQ corresponding to the maximum likelihood parameter set.

The resulting likelihoods were sorted into descending order, along with the associated parameter vectors, and effectively converted into a lookup table for a sample-based cumulative posterior pdf. This allows straightforward random sampling from the posterior. Approximately 10,000 parameter vectors have non-negligible weight in the posterior.

Percentiles of predicted daily detected local cases, based on an ensemble of 200 DCM trajectories using parameter sets drawn randomly from the posterior, are compared with the observations in Fig. 1. The fit is generally, good but observe that the model does not adequately capture the steep rise in reported cases in the state of Victoria in late June – early July 2020. An equivalent ensemble for the SCM shows additional scatter due to stochastic effects (Fig. 2). For the SCM, the observed steep increase in detected local cases in late June – early July 2020 does lie within the inter-quartile credibility interval.

Comparisons of SCM posterior ensemble predictions of cumulative deaths and cumulative detected recoveries are shown in Fig. 3, 4. As indicated above, the choice of parameters T_D_, P_M_ and P_D_ provide good approximations to the timing and magnitude of observed cumulative deaths through the ‘first wave’. The model arguably predicts deaths too early in the current ‘second wave’ in the state of Victoria. We observe the outbreak in Victoria that began in June 2020 started in a relatively younger demographic of workers and students, and only later spread into aged care homes in July 2020 which has resulted in additional deaths.

The choice of T_R_ and P_R_ results concur well with the period between predicted and observed recoveries in detected cases (Fig. 4), given some obvious reporting anomalies in the observations. The median model prediction slightly over predicts recoveries, but given the good fit to cases, it seems possible this reflects incomplete official reporting of recoveries.

The marginal pdfs for the parameters, calculated from the posterior weighted sample, are plotted as histograms in Fig. 5. The parameters fall into three groups with respect to the information provided by the observations. The parameters G0 and G_LD_ are highly informed, with posterior values restricted to a narrow range. The parameters RSD, P_U_ and P_L_ are moderately informed, with RSD biased to high values between 0.8 and 1, P_U_ to high values from 0.4 to 0.6 and P_L_ biased towards low values from 0.1 to 0.2. The remaining parameters seem to be effectively uninformed, with little difference between prior and posterior.

The posterior distribution carries additional information in the form of correlations among parameters (Fig. 6). A strong negative correlation between G_LD_ and P_U_ suggests the model is relying partly on contact tracing to bring about the steep decline in local cases in March-April 2020. High levels of uncooperative behaviour weaken contact tracing, and more effective social distancing (lower G_LD_) is then required to match observations. There are also moderate negative correlations between RSD and P_U_, and between G0, G_LD_ and P_L_, with similar explanations. A weaker positive correlation between G0 and P_T_ suggests contact tracing reduces net transmissions prior to implementation of social distancing. The positive correlation between P_U_ and P_L_ seems more likely to be an indirect result of their mutual strong negative correlations with transmission coefficients.

The DCM provides a relatively tight fit to observations despite the majority of the parameters being poorly informed. We posit this may be because of trade-offs among parameters, reflected in the correlation structure, noting that some poorly informed parameters such as P_A_ and F_A_ are weakly correlated with other parameters (Fig. 6). We contend that scenario ensembles based on random samples from this posterior provide a realistic picture of model uncertainty given current knowledge.

**Table 6.**
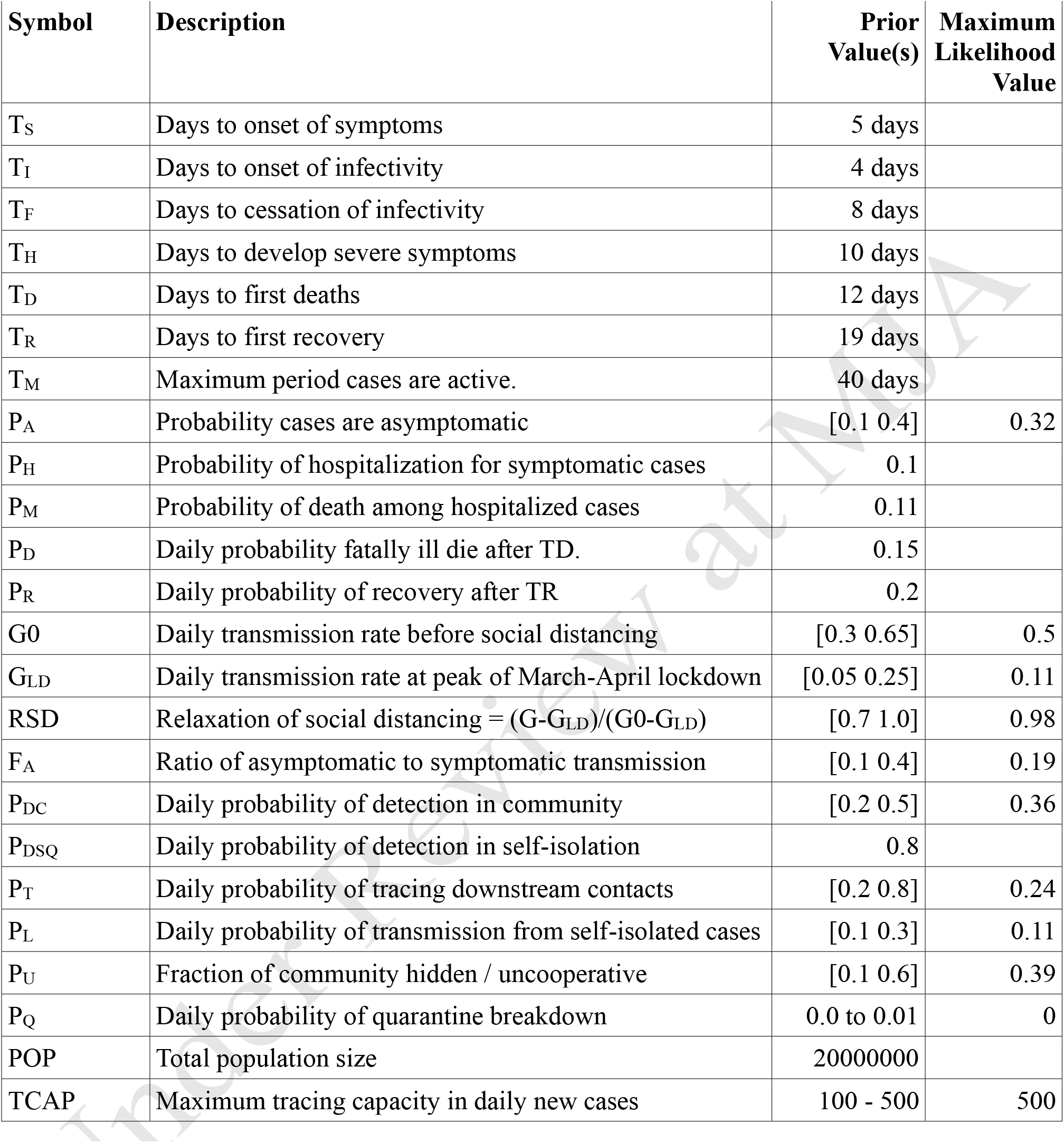
Model parameters, prior values, and prior ranges and maximum likelihood values for those parameters subject to Bayesian inference.

**Fig. 1.**
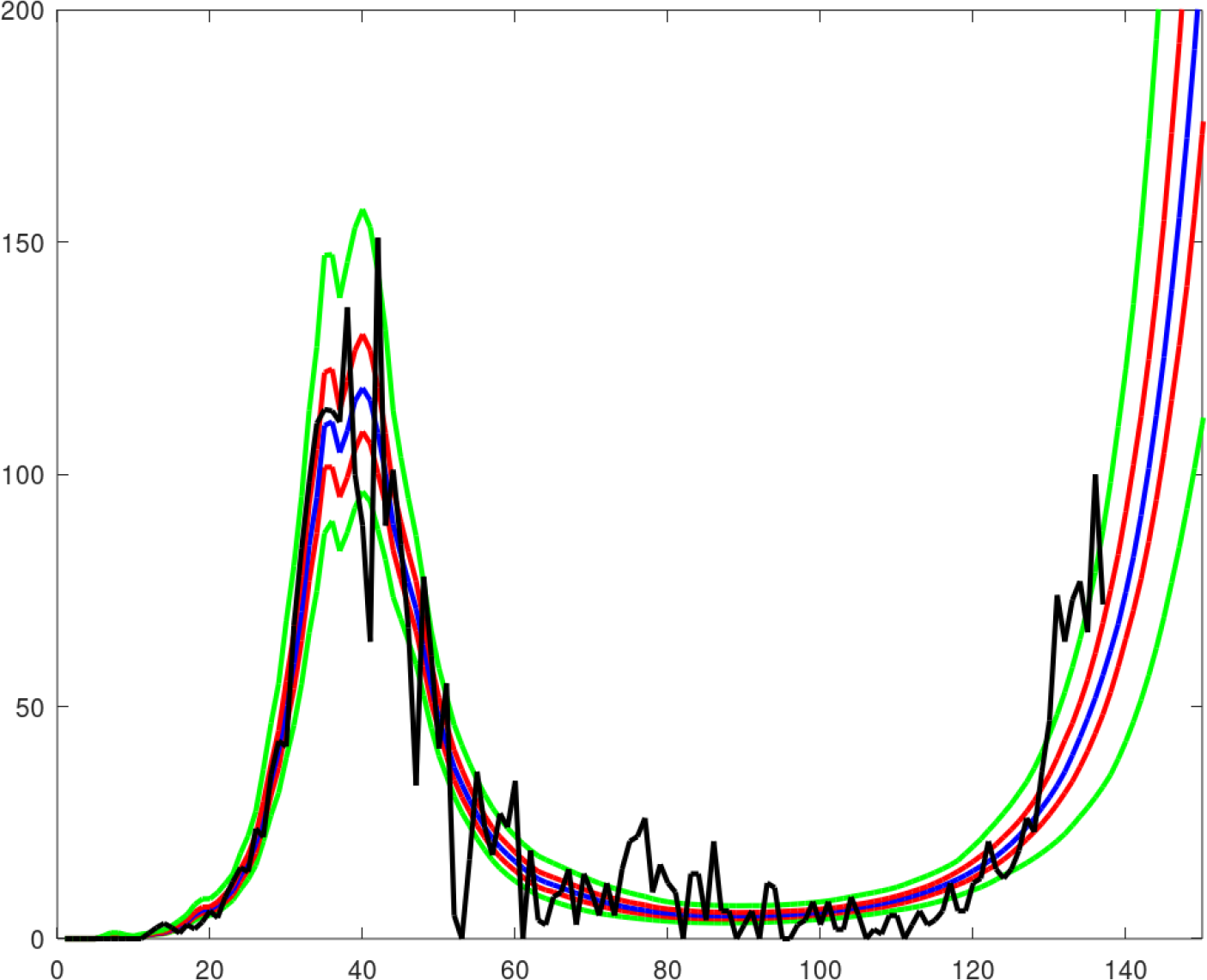
Percentiles [2.5% 25% 50% 75% 97.5%] of detected local Australian cases for a posterior ensemble of trajectories from the DCM, with observations in black. X-axis shows days starting 20_th_ February 2020.

**Fig. 2.**
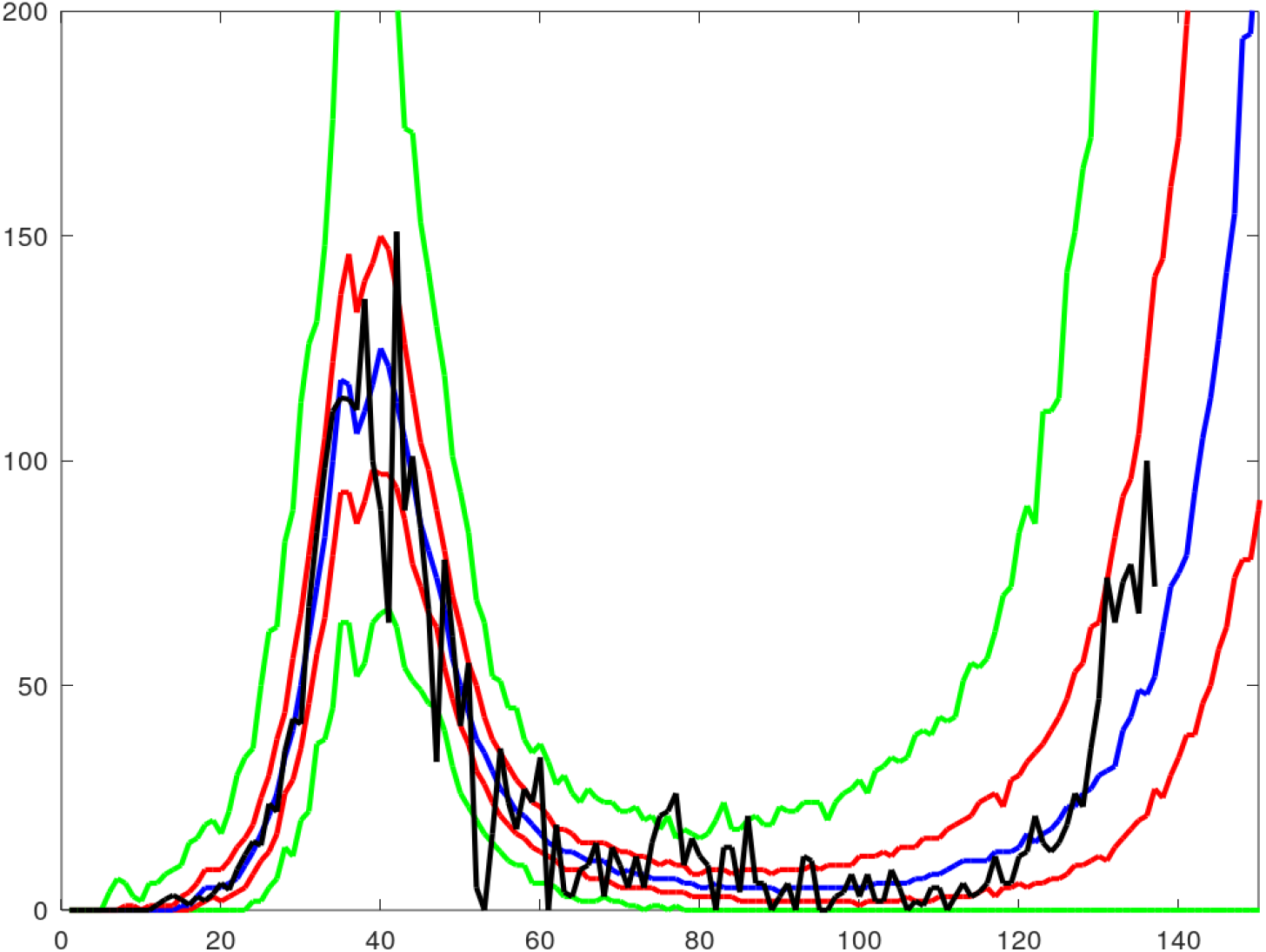
Percentiles [2.5% 25% 50% 75% 97.5%] of detected local Australian cases for a posterior ensemble of trajectories from the SCM, with observations in black. X-axis shows days starting 20^th^ February 2020.

**Fig. 3.**
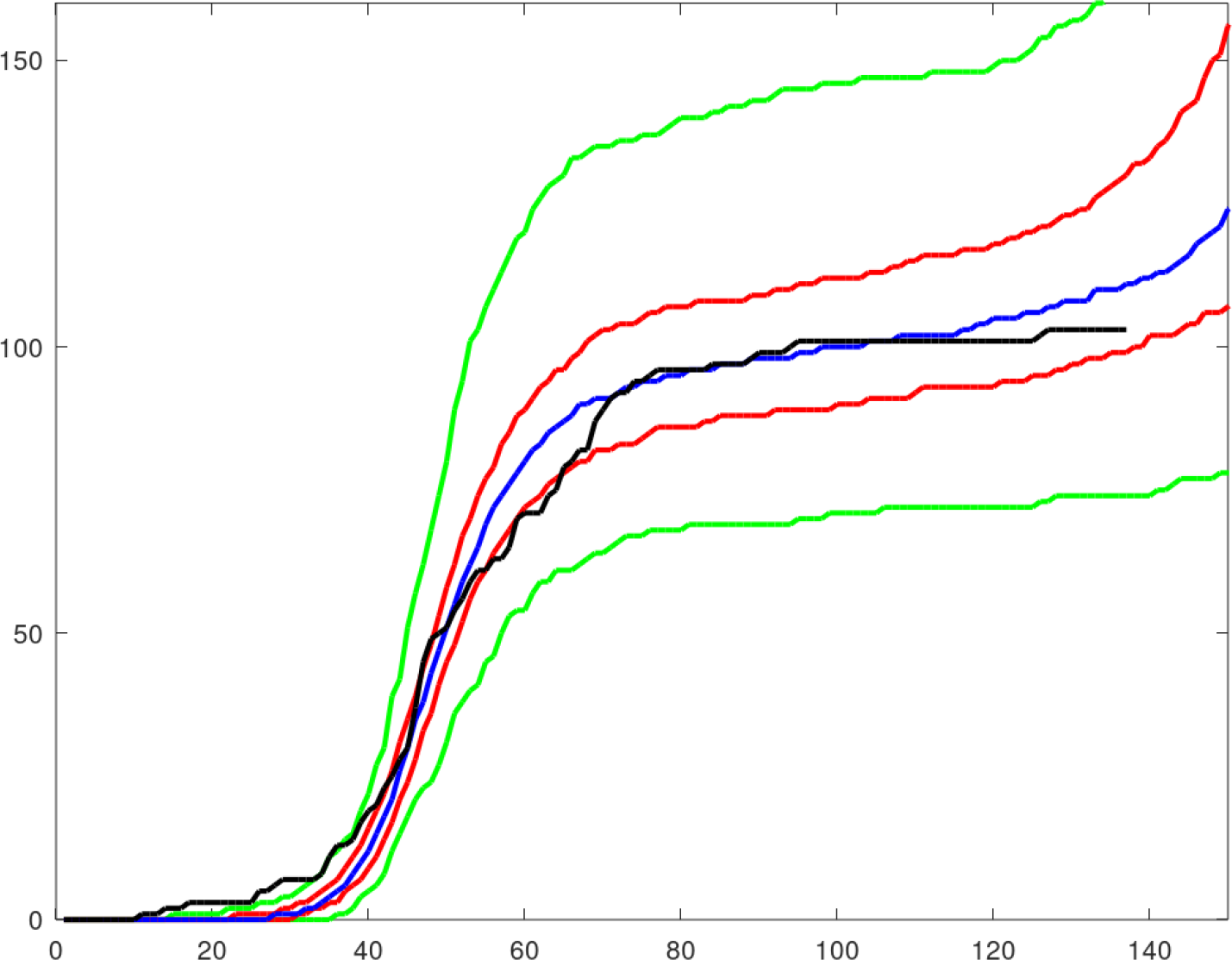
Percentiles [2.5% 25% 50% 75% 97.5%] of cumulative deaths for posterior ensemble from the SCM. Observed cumulative deaths in black. X-axis represents days since 20^th^ February 2020.

**Fig. 4.**
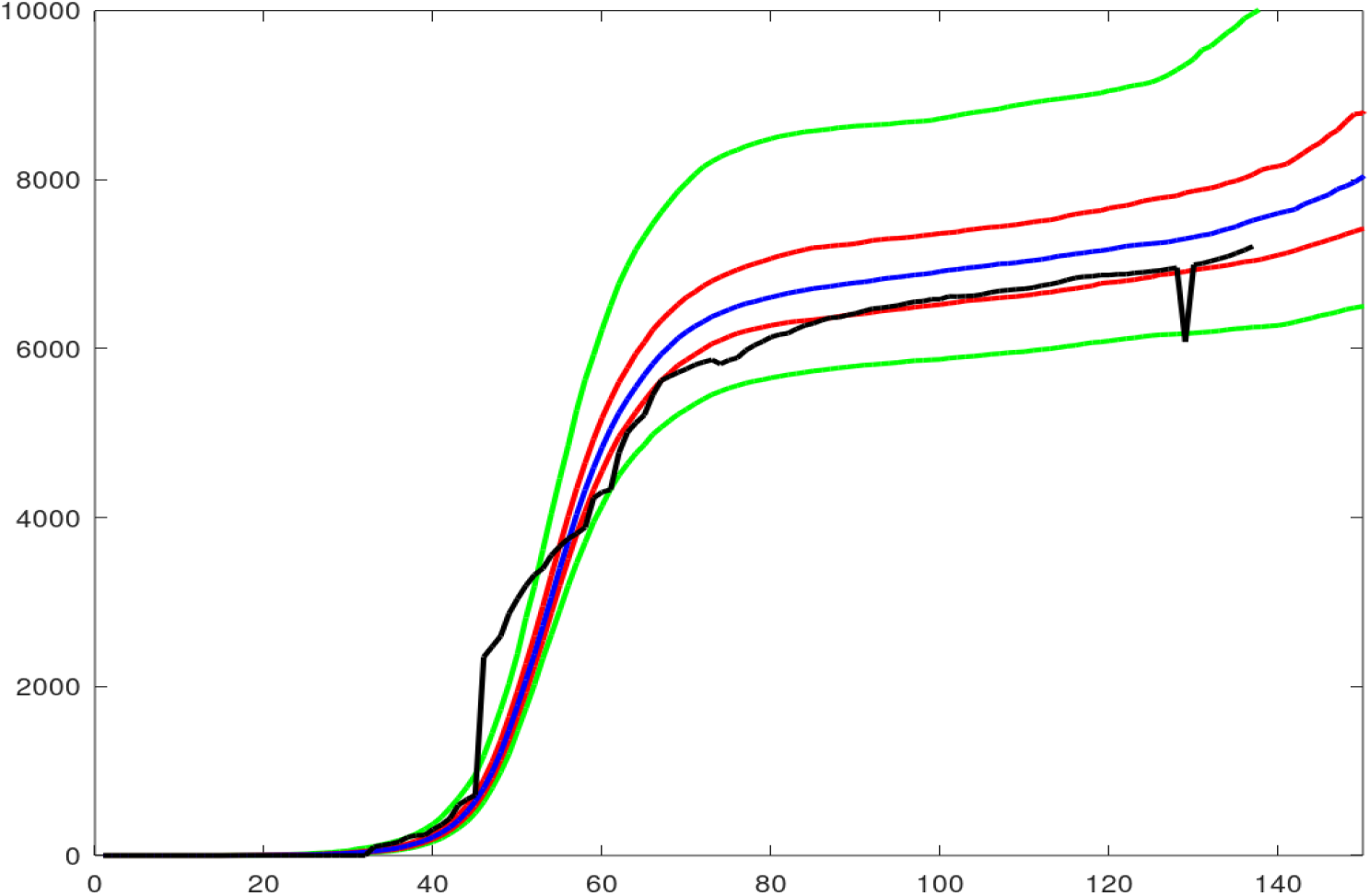
Percentiles [2.5% 25% 50% 75% 97.5%] of cumulative detected recoveries for posterior ensemble from the SCM. Observed cumulative detected recoveries in black. X-axis represents days since 20^th^ February 2020.

**Fig 5A.**
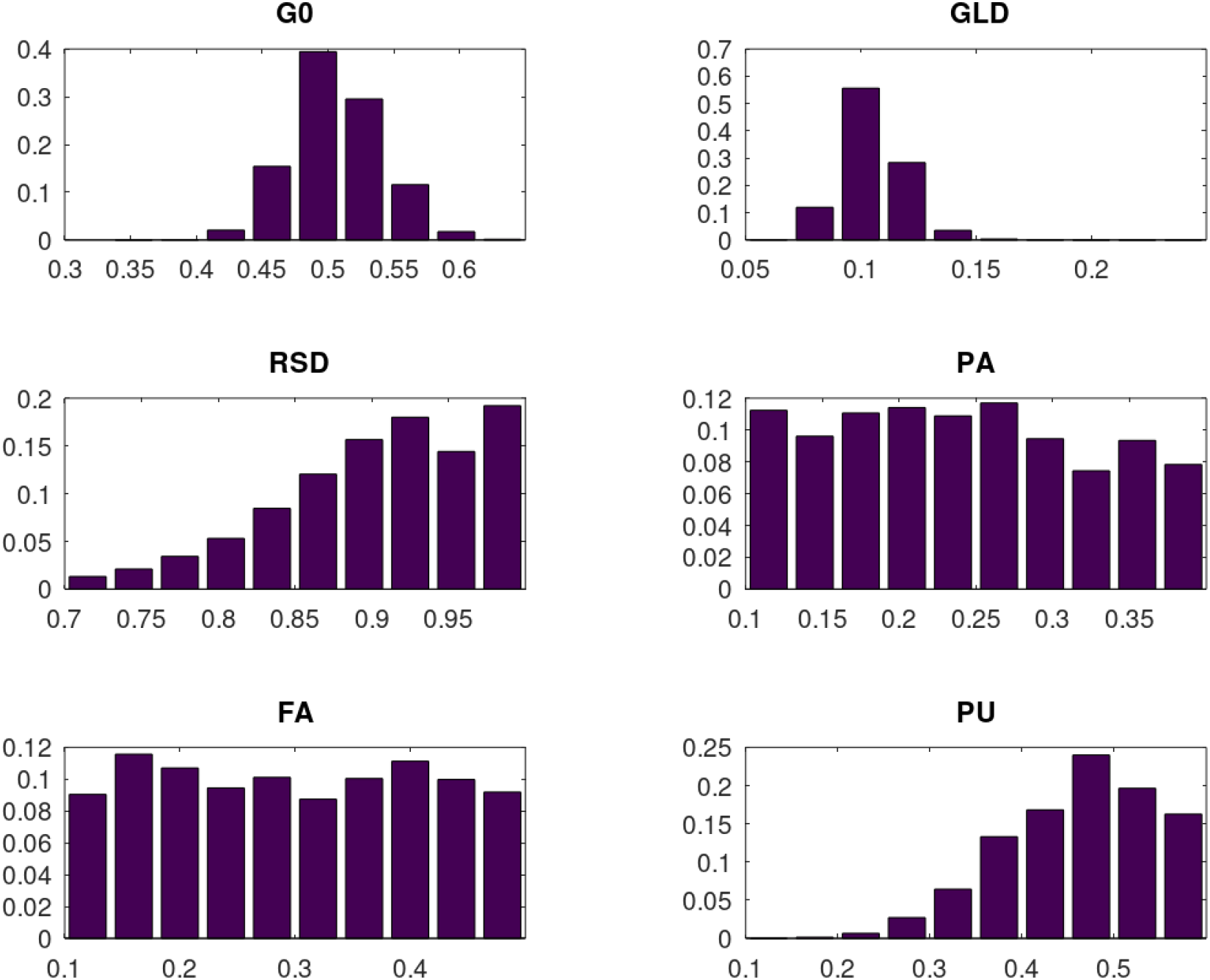
Marginal posterior pdfs for parameters subject to Bayesian inference. (G0, GLD – Initial and lockdown transmission rates, RSD – relaxation of social distancing by 5 July 2020 PA – proportion asymptomatic, FA – relative transmission of persons who are asymptomatic, PU – proportion unco-operative.)

**Fig 5B.**
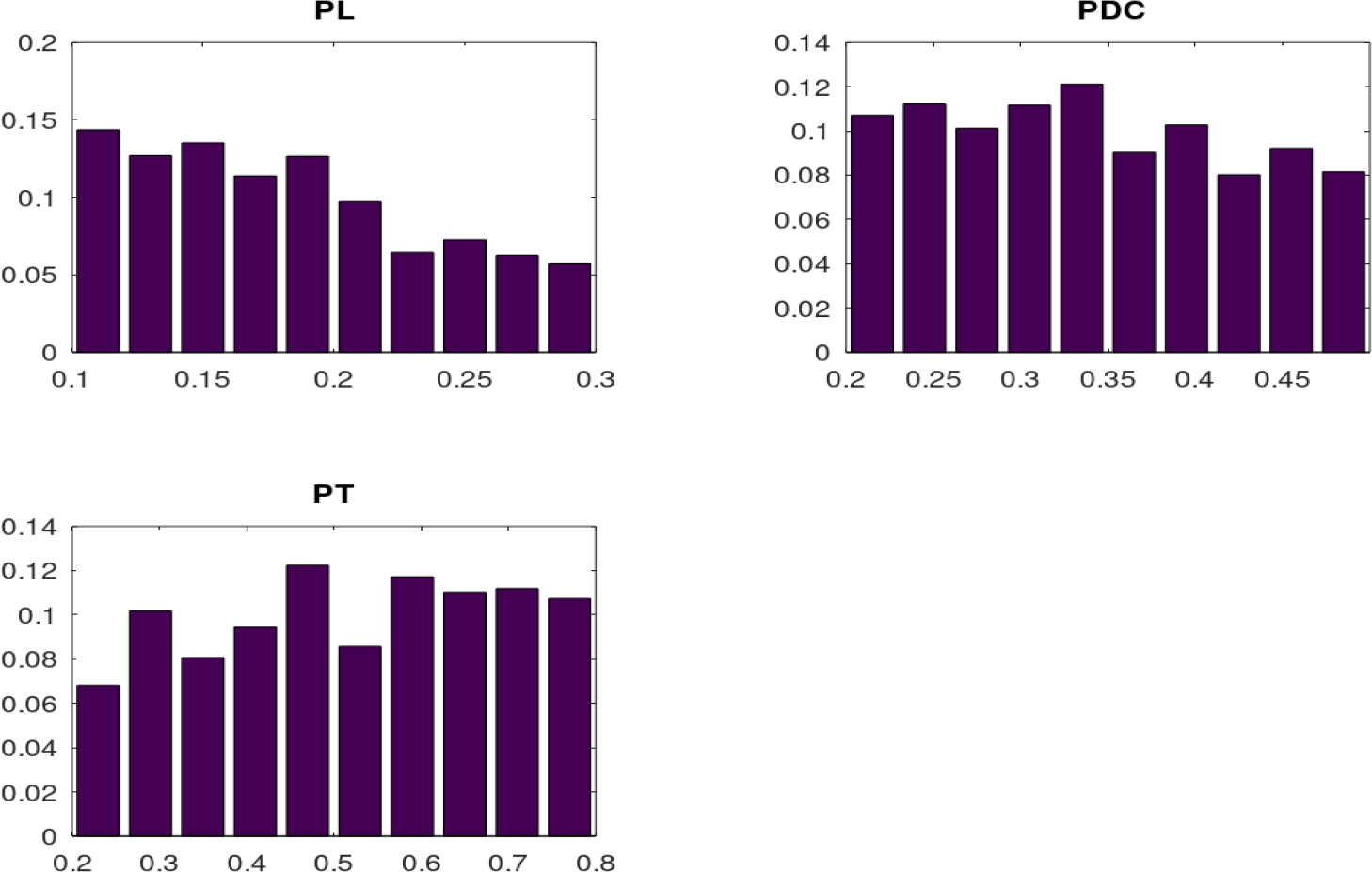
Marginal posterior pdfs for parameters subject to Bayesian inference. (PL – probability of violation of self-quarantine, PDC – probability of detection in community, PT – probability of downstream tracing.)

**Fig. 6.**
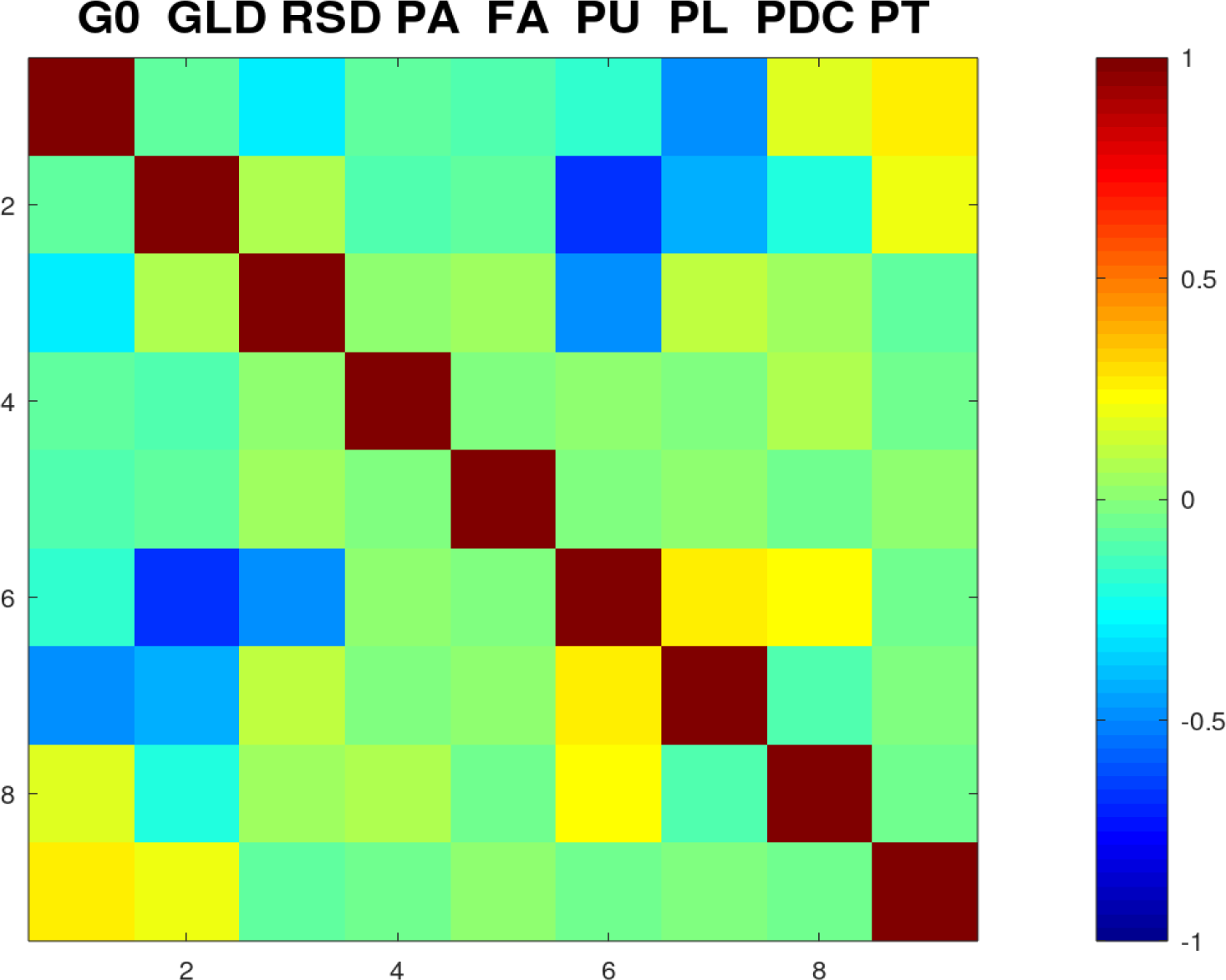
Posterior correlation structure among parameters subject to Bayesian inference. (G0, GLD – Initial and lockdown transmission rates, RSD – relaxation of social distancing by 5 July, 2020 PA – proportion asymptomatic, FA – relative transmission of persons who are asymptomatic, PU – proportion unco-operative. PL – probability of violation of self-quarantine, PDC – probability of detection in community, PT – probability of downstream tracing.)

